# Scalable deep-learning-based inference of time-varying transmission dynamics from outbreak phylogenies

**DOI:** 10.64898/2026.05.07.26352673

**Authors:** Ruopeng Xie, Anna Zhukova, Pablo G. Peña, Guillermo Iglesias, Shu Hu, Jiawei Wang, Tim K. Tsang, Vijaykrishna Dhanasekaran, Moritz U. G. Kraemer, Oliver G. Pybus, Olivier Gascuel

## Abstract

Infectious disease dynamics can be inferred from pathogen genomic data using phylodynamic methods, but the applicability of many such approaches to large data sets is constrained by computational cost. Recent deep-learning approaches to phylodynamics have improved scalability, yet challenges remain when genetic divergence is limited during fast spreading outbreaks. To address this, we use pathogen-specific models to show that deep-learning models trained on outbreak-like phylogenies can accurately estimate the reproductive number (*R*) when both the birth–death model and the expected phylogenetic resolution are matched to the target pathogen, highlighting the importance of realistic training conditions. Focusing on three major respiratory pathogens of public health importance (SARS-CoV-2, seasonal human influenza virus, and respiratory syncytial virus (RSV)), we introduce *PhyloRt*, a scalable framework for estimating the time-varying reproductive number (*R_t_*) from large outbreak phylogenies. *PhyloRt* decomposes large trees into overlapping subtrees and applies a hierarchical deep-learning-based inference strategy to classify subtrees as exhibiting constant or time-varying reproduction numbers, enabling identifiable and computationally efficient estimation of *R_t_* as a piecewise-constant trajectory through time. Applications to SARS-CoV-2 and influenza outbreaks show that *PhyloRt* recovers transmission dynamics consistent with estimates derived from mathematical epidemiological and Bayesian phylodynamic analyses. Our work enables scalable and rapid estimation of time-varying transmission dynamics from very large-scale outbreak genomic data sets, supporting real-time genomic epidemiology of emerging pathogens.

**Significance:** Estimating changes in transmission dynamics over time is important for responding to infectious disease outbreaks. Current methods mostly rely on reported case data from epidemiological surveillance, which can be biased or incomplete due to variable testing capabilities, particularly in resource-limited settings. A complementary approach is to use viral genomes as an alternative data source. However, inferences from genomic data can be computationally intensive and have mainly been applied retrospectively. We present PhyloRt, a scalable deep-learning–based phylodynamic framework that enables fast inference of the time-varying reproductive number (Rt) from large outbreak phylogenies. Our approach is widely applicable and provides a practical approach to monitoring epidemic dynamics, complementing traditional surveillance and supporting timely public health decision-making.

## Introduction

Reconstructing epidemic dynamics in real-time has become increasingly important for infectious disease management, as demonstrated during the COVID-19 pandemic (e.g.(1)). Traditional epidemiological methods have relied on reported case data, which can be biased or incomplete due to changes in testing policies and capacity, particularly in resource-limited settings (2). To address these issues, methods that use alternative data sources have been developed (e.g., social media (3) or wastewater data (4, 5)), with the aim of improving the timeliness and accuracy of estimated transmission parameters. More recently, large scale pathogen sequencing surveillance efforts have enabled the tracking of infectious disease spread using genomic data (6).

Phylodynamics is an interdisciplinary field that integrates evolutionary and epidemiological processes and offers unique perspectives on epidemic history, geographic spread, disease dynamics (6–8). Its strength lies in complementing traditional epidemiological approaches with genomic data, particularly when conventional data is sparse or absent (9). Moreover, by reconstructing transmission lineages, phylodynamics can distinguish between local and imported cases (10). Despite its demonstrated impact, state-of-the-art phylodynamic methods remain computationally intensive due to the vast parameter space they need to explore during inference (11), limiting their use for real-time epidemiological modelling when data sets are large.

In response, significant efforts have been made to optimize algorithms and analytical pipelines, enhancing computational efficiency (6, 12–14). Notably, PhyloDeep (14) integrates deep learning with phylodynamics by training neural networks to predict epidemiological parameters from simulated trees generated under birth–death (BD) models, rather than maximizing the likelihood of observed data under those models. This approach, which is conceptually related to approximate Bayesian computation (ABC) (15), enables rapid parameter estimation from very large phylogenetic trees within seconds. PhyloCNN (16) builds on this by encoding local tree features into a convolutional neural network, achieving accurate inference with training datasets one to two orders of magnitude smaller than those used previously. However, challenges remain when pathogen genetic divergence is limited during well sampled outbreaks. Our recent prior study (17) showed that PhyloDeep models, trained on high-resolution simulated trees from birth-death models, can underperform on outbreak datasets such as COVID-19. However, retraining on pathogen-specific poorly resolved, realistic trees restores accuracy for estimating the reproductive number (*R*) (17), offering a promising avenue for tracking epidemic dynamics using sequence data.

Here, we introduce *PhyloRt*, a scalable phylodynamic framework that combines deep learning with hierarchical subtree-based inference to estimate the time-varying reproductive number (*R_t_*) from large outbreak phylogenies. We first examine how different birth–death model formulations affect parameter inference when focusing on *R* estimation. We then apply this framework to three major respiratory pathogens of continued public health importance: SARS-CoV-2, seasonal human influenza virus, and respiratory syncytial virus (RSV). By decomposing large phylogenies into overlapping subtrees and aggregating subtree-level predictions, *PhyloRt* enables robust estimation of temporal transmission dynamics while remaining computationally efficient on large genomic datasets. We explore the performance of our method on simulated and real-world outbreak data.

## Results

### Pathogen-specific neural network models

We simulated phylogenetic trees using several types of BD model and transformed them into realistic, pathogen-specific phylogenies by assigning genetic distances to branches using a Poisson process, collapsing zero-length branches, randomly resolving polytomies, and re-dating trees using LSD2 (18). These steps aim to mimic the limited genetic resolution of outbreak data (see Methods for details, SI Appendix, Tables S1-2). Neural network models were then trained using the PhyloCNN framework (16) (Fig. 1A). Because superspreading parameters cannot be inferred reliably from such poorly resolved trees (17), we first assessed whether the basic BD model is sufficient when focusing solely on estimating *R*. Using SARS-CoV-2 as an example across four BD model versions (Fig. 1B; SI Appendix, Figs. S1-2 and S.1), we found that accurate inference requires matching the BD model used for training to that underlying the data, as mismatched models lead to systematic bias. In particular, models trained on simpler BD formulations tend to underestimate *R* when applied to more complex transmission processes; for example, applying a BD-trained model to trees simulated under a BD model with exposed–infectious classes and superspreading (BDEISS) yielded a mean relative error (MRE) of 0.325 and a mean relative bias (MRB) of −0.318 (SI Appendix, Figs. S1-2).

**Fig. 1.**
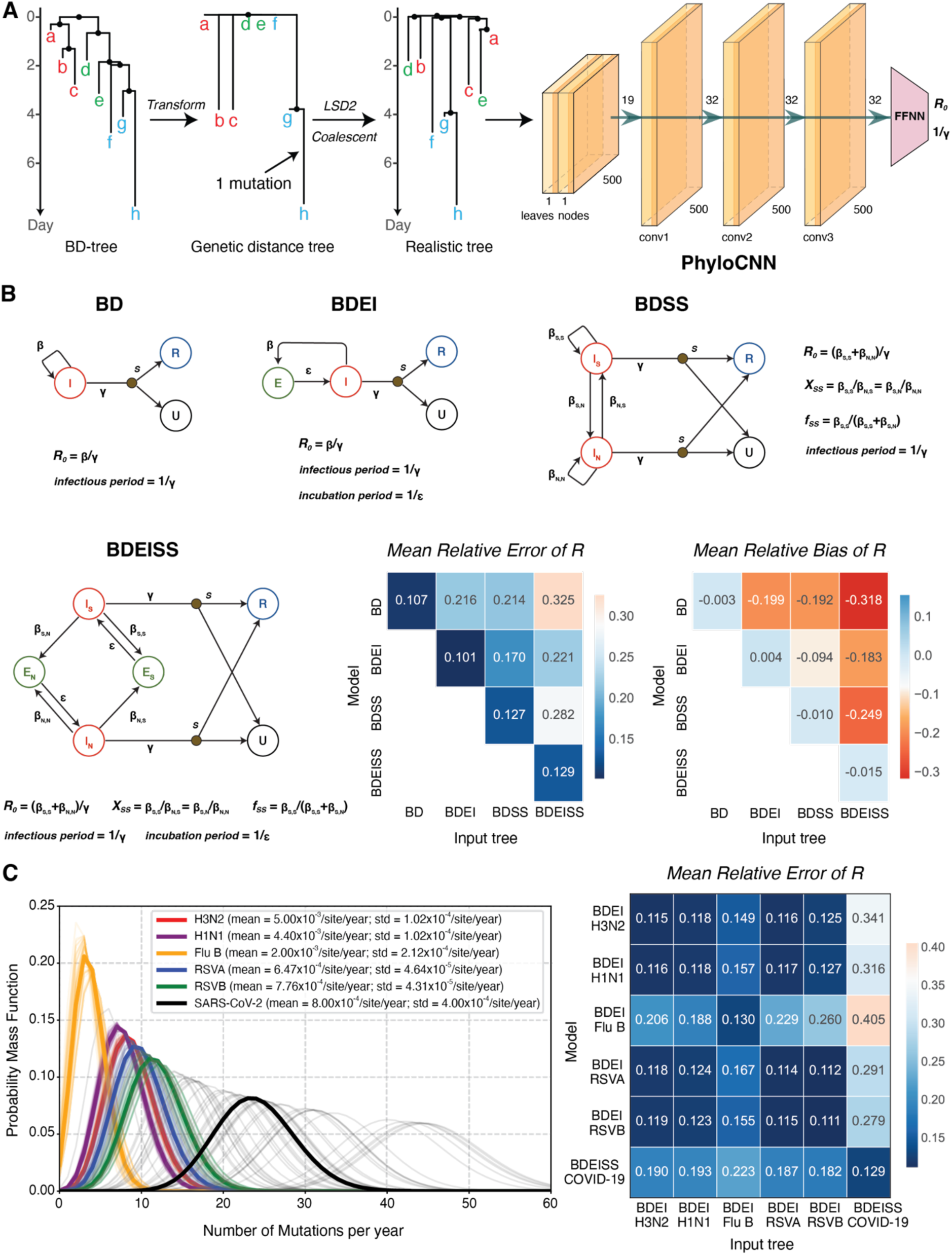
Pathogen-specific neural network models for phylodynamic inference from realistic phylogenies. **(A)** Schematic of the workflow under constant transmission rates (tree size = 200-500 tips). High resolution phylogenetic trees were simulated using birth–death models (BD-tree) and transformed into genetic distance trees, with branches containing zero mutations collapsed. Any resulting polytomies were then randomly resolved using a coalescent model, after which the trees were converted to time-scaled trees using LSD2 (18) to obtain realistic, binary time-scaled trees. These trees were used as inputs to the PhyloCNN framework, which extracts features from leaves and nodes through convolutional layers (conv1-to-3) and predicts epidemiological parameters using a feed-forward neural network (FFNN). **(B)** Birth–death model variants used for simulation: basic birth–death (BD), birth–death with exposed and infectious classes (BDEI), birth–death with superspreading (BDSS), and birth–death with both exposed–infectious classes and superspreading (BDEISS). Parameters include *R₀*, infectious period (1/γ), incubation period (1/ε), proportion of superspreaders (*fₛₛ*), and relative transmission rates of superspreaders to normal spreaders (*Xₛₛ*). These birth-death model illustrations are adapted from PhyloDeep (14). Heatmaps show mean relative error (MRE) and mean relative bias (MRB) of *R₀* estimates for models trained and tested on all combinations of realistic trees characterised by SARS-CoV-2 sequence length and divergence rates under each BD model type. **(C)** Left: Divergence rate distributions used for each respiratory virus data set. Standard deviations were derived from published 95% confidence intervals under a normal approximation (SI Appendix, Table S2). For influenza viruses, simulations were based on the hemagglutinin (HA) segment only. Right: Heatmap shows MRE of *R₀* estimates for pathogen-specific models trained on realistic trees under matched birth–death models for each pathogen, evaluated across all model–input tree combinations.

Next, we focused on three major respiratory viruses: SARS-CoV-2, human seasonal influenza (influenza A subtypes H3N2 and H1N1pdm09; influenza B lineages Victoria and Yamagata, combined due to their similar substitution rates (19)), and RSV (subtypes A and B). For each pathogen, we simulated high-resolution phylogenies using a biologically realistic birth–death model matched to its transmission dynamics (BDEISS for COVID-19 (20); birth–death with exposed and infectious classes (BDEI) for influenza (21, 22) and RSV (23)). These phylogenies were transformed to realistic trees according to pathogen-specific genome lengths and substitution rates, to match the resolution typically observed in observed outbreak datasets. We compared performance across all model combinations (Fig. 1C). Notably, models for influenza A (BDEI-H3N2, BDEI-H1N1) and RSV (BDEI-RSVA, BDEI-RSVB) demonstrated mutual compatibility, with less than 1% increase in MRE, whereas influenza B and SARS-CoV-2 required models that matched their pathogen-specific biology (BDEI-FluB and BDEISS-COVID-19). This compatibility likely reflects similar levels of phylogenetic resolution, characterized by overlapping rates of molecular divergence that are influenced by genome length and clock rate (Fig. 1C).

### Hierarchical subtree-based inference enables estimation of *R_t_*

Transmission dynamics vary with changes in public health interventions, population immunity, and behavioural patterns. We introduce *PhyloRt*, a hierarchical inference framework based on scalable subtree decomposition that enables identifiable reconstruction of *R_t_* using a piecewise-constant formulation (24). Neural network models were trained within the PhyloCNN framework using 50-400 tip subtrees, pruned from 10,000 large phylogenetic trees, each containing 1,000 to 10,000 tips, simulated under an epidemiological scenario in which the reproductive number changes at a single time point (Fig. 2A; Methods). This training procedure yielded three task-specific models: a classification model for detecting the presence of a change point, a regression model for estimating a single reproductive number (*R_NC_*) from subtrees without a change point, and a regression model for estimating the reproductive number before (*R_C1_*) and after (*R_C2_*) the change point, together with the change-point position from subtrees containing a change point (Fig. 2A). During inference, an input tree is decomposed into overlapping subtrees, the change-point probability is first estimated for each subtree, and parameter inference is then performed, conditional on whether the subtree is classified as containing a change point or not. Conservative probability thresholds (<0.1 and >0.9) were used to achieve positive predictive values (PPV) and negative predictive values (NPV) exceeding 90% (SI Appendix, Fig. S4 and Tables S3-4). Subtree-level predictions were subsequently aggregated across time to recover a piecewise-constant estimate of *R_t_* using 14-day time windows (Figs. 2B-C).

**Fig. 2.**
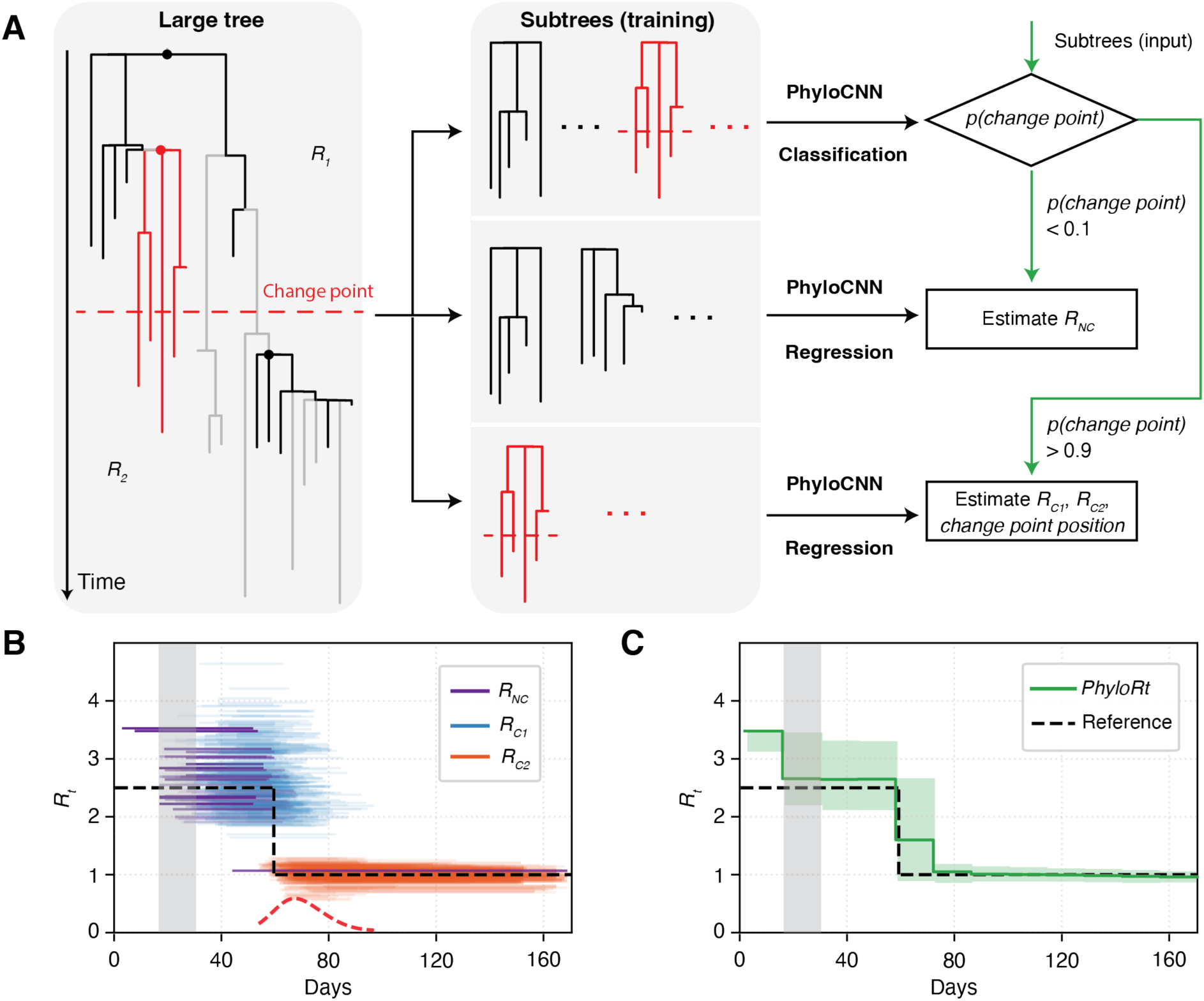
Overview of the *PhyloRt* framework for *R_t_* estimation. (A) Subtree-based inference workflow in *PhyloRt*. **A** total of 10,000 large phylogenetic trees were simulated, each comprising 1,000–10,000 tips and a single change point that separates two reproductive-number regimes (*R_1_* and *R_2_*). These trees were subsequently pruned into overlapping subtrees of predefined sizes. The figure provides a schematic illustration of a large tree and three example subtrees (red subtrees contain a breakpoint, whilst black subtrees do not). These subtrees are used to train classification and regression models using PhyloCNN. During inference, subtrees extracted from the input tree are first evaluated using the classification model to estimate the probability of a change point. Subtrees with low change-point probability (p < 0.1) are analysed using a regression model to estimate a single reproductive number (*R_NC_*), whereas subtrees with high change-point probability (p > 0.9) are analysed using regression models to estimate the reproductive number before (*R_C1_*) and after (*R_C2_*) the change point, together with the change-point position**. (B)** Subtree-level *R* estimates through time. Each line represents an estimate inferred from an individual subtree, spanning from the subtree root age to its most recent sampling date and coloured according to the inferred transmission phase (*R_NC_*, *R_C1_*, or *R_C2_*). The dashed red curve shows a kernel-smoothed density of inferred change-point times across subtrees, normalized and scaled for visualization. The dashed black line indicates the true underlying *R*. The shaded vertical region highlights an example time bin used for aggregating subtree-level estimates. **(C)** *R_t_* inferred by *PhyloRt*. Subtree-level reproductive number estimates are aggregated within time bins using a weighted mean (see Methods for details) to produce a piecewise-constant estimate of *R_t_* (green line), with shaded bands indicating the interquartile range (IQR; 25th–75th percentiles) of subtree-level predictions. The shaded vertical region corresponds to the same example time bin shown in panel B.

### Subtree size affects stability and temporal resolution

We evaluated *PhyloRt* under four transmission scenarios (constant *R_t_*, increase in *R_t_*, decrease in *R_t_*, and alternating *R_t_* values) using simulated trees with 2,000 to 3,000 tips, and compared performance against a baseline direct-averaging approach that does not perform change-point detection and instead infers a single reproductive number (*R_NC_)* per subtree under a constant-*R* assumption. Analyses focused on the H3N2, influenza B, and SARS-CoV-2 data sets (Fig. 3, SI Appendix, Figs. S5-10 and Table S5), as models for RSV and H1N1pdm09 showed strong compatibility with those for H3N2 (Fig. 1C). Throughout, subtree-based inference was performed using subtree tip sizes of 50, 100, 200, and 400.

**Fig. 3.**
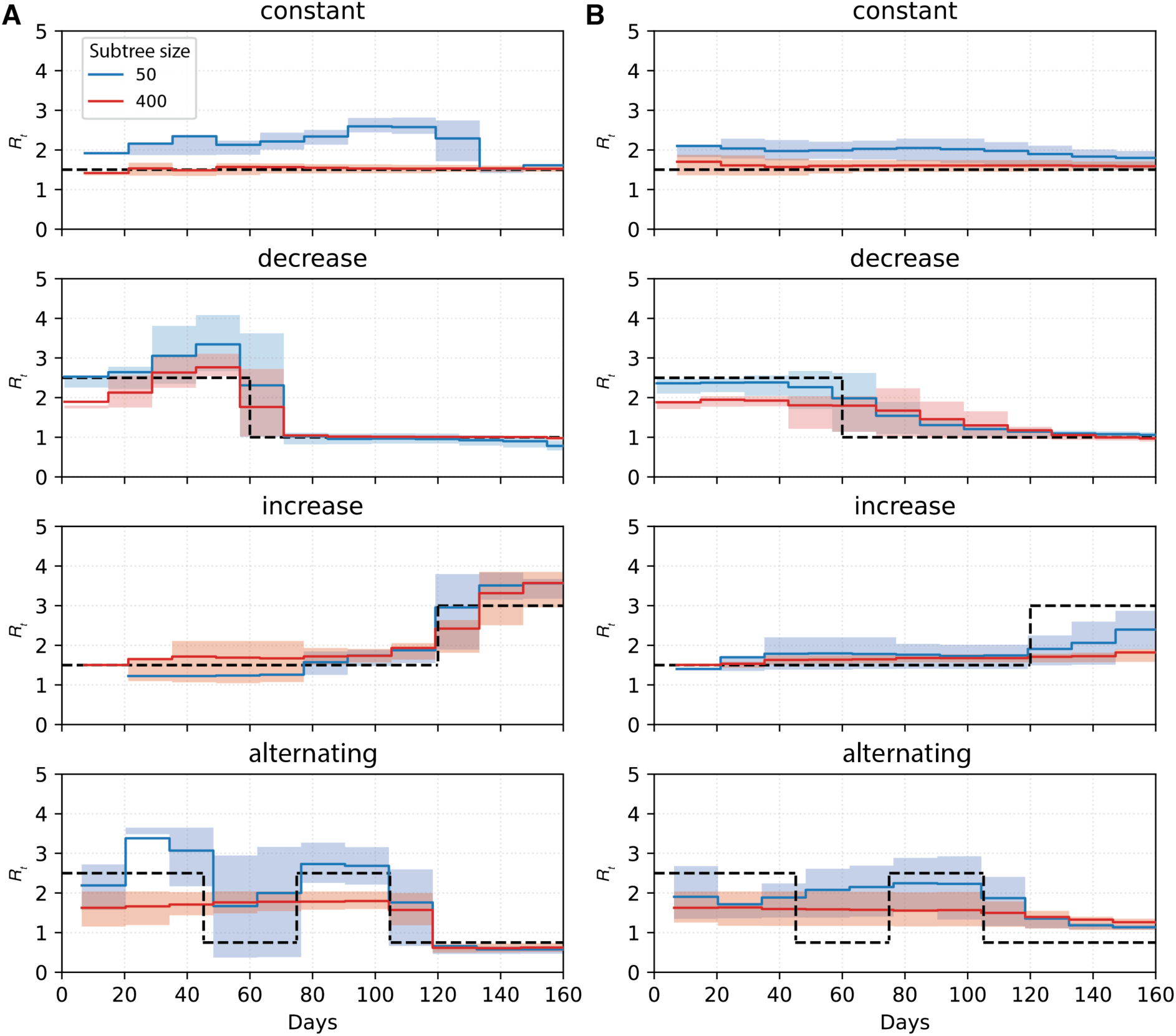
Comparison of (A) hierarchical and (B) baseline direct-averaging subtree-based inference for estimating *R_t_*in three independently simulated H3N2 trees with 2,000–3,000 tips across four transmission scenarios. Colours indicate subtree size (number of tips). Solid lines show the estimated mean *R_t_*, with shaded bands representing the IQR. Dashed black lines show the true underlying trajectory of *R* through time. Performance of the neural network models across subtree sizes is shown in SI Appendix, Fig. S4.

Across all pathogens and scenarios, the hierarchical subtree-based approach more accurately captured changes in *R_t_* than direct averaging. Performance improved systematically with increasing subtree tip size, with subtrees of 200 and 400 tips yielding more robust and stable estimates, consistent with improved neural network performance at larger tip sizes (SI Appendix; Fig. S4) and with our previous findings (14). For example, under hierarchical inference across constant, decreasing, and increasing *R* scenarios, for subtrees of 200 and 400 tips, time-normalized mean absolute error (MAE) remained below 0.4 for H3N2, below 0.6 for influenza B, and below 0.3 for SARS-CoV-2 (SI Appendix, Figs. S5-10 and Table S5). However, larger subtrees also tended to smooth short-term fluctuations, particularly in the alternating *R* scenario, whereas smaller subtrees (50 tips) were better at capturing rapid temporal changes. For example, MAE reached 0.389 for H3N2 and 0.392 for SARS-CoV-2 using 100-tip subtrees, and 0.512 for influenza B using 50-tip subtrees.

As the size of the full tree increased to 5,000–10,000 tips, larger subtrees (200 and 400 tips) were also able to recover short-term temporal signals in the alternating *R* scenario when a recency-based weighting was applied (see Methods; SI Appendix, Figs. S11-12 and Table S6), achieving MAE of 0.307 for 200-tip subtrees. However, both small subtrees and recency-based weighting increase sensitivity to short-term changes and therefore share a common trade-off in reduced estimation stability. Importantly, *PhyloRt* remained computationally efficient, with inference on each phylogeny (containing up to 10,000 tips) completing in under 10 minutes on a MacBook Air M4.

Overall performance was highest for H3N2, followed by SARS-CoV-2 and influenza B, indicating that differences in birth–death model complexity and genetic diversity affect the amount of information retained in realistic phylogenies and the extent to which this information can be leveraged by neural network models, thereby influencing the recovery of temporal dynamics.

### Application of *PhyloRt* to real outbreak datasets

We applied *PhyloRt* to two real outbreak datasets: SARS-CoV-2 in the United Kingdom (UK) in early 2020 (6) and influenza B in China during 2020–2021 (25). The former is characterized by extensive surveillance data and large-scale, well-represented genomic sampling, thereby enabling direct comparison of phylodynamic inference with incidence-based estimates, whereas the latter dataset is of moderate size, allowing direct comparison with Bayesian phylodynamic inference using BEAST2 (26).

### SARS-CoV-2 in the United Kingdom

The United Kingdom experienced one of the largest SARS-CoV-2 epidemics worldwide in early 2020, with rapid case growth in March, peaking in April. Widespread viral importation during this period produced >1,000 distinct transmission lineages (6). In order to focus on sustained local transmission, we analyzed the 50 largest UK local transmission lineages (sampled 2 Feb–17 Jun 2020; lineage size = 86–1,865 tips), comprising 13,883 genome sequences in total. Each lineage was analyzed independently using *PhyloRt*, and subtree-level estimates were aggregated across all lineages over time to reconstruct final *R_t_*.

Mathematical epidemiological estimates indicated a pre-intervention *R_t_* = 3.82 (95% CI: 3.36–4.36), falling to *R_t_* = 0.65 (95% CI: 0.52–0.78) after the national lockdown on 24 March 2020 (27). *PhyloRt* recovered the same overall dynamic. Estimates based on small subtrees (50 tips) captured the early and late *R* values, with a mean inferred change-point date of 26 March 2020 (95% CI: 8 March–12 April 2020), but exhibited a relatively gradual transition, contrasting with the very sharp change observed in mathematical epidemiological estimates (Figs. 4A-B). This reflects the fact that the latter estimate was based on an approach that modelled *R_t_* as a piecewise-constant function that changes only at intervention dates (27). Larger subtrees (200–400 tips) reproduced the same qualitative pattern but yielded lower *R_t_* estimates prior to mid-March, likely due to temporal averaging over longer subtree spans that smooths over short-term increases (SI Appendix, Fig. S15).

**Fig. 4.**
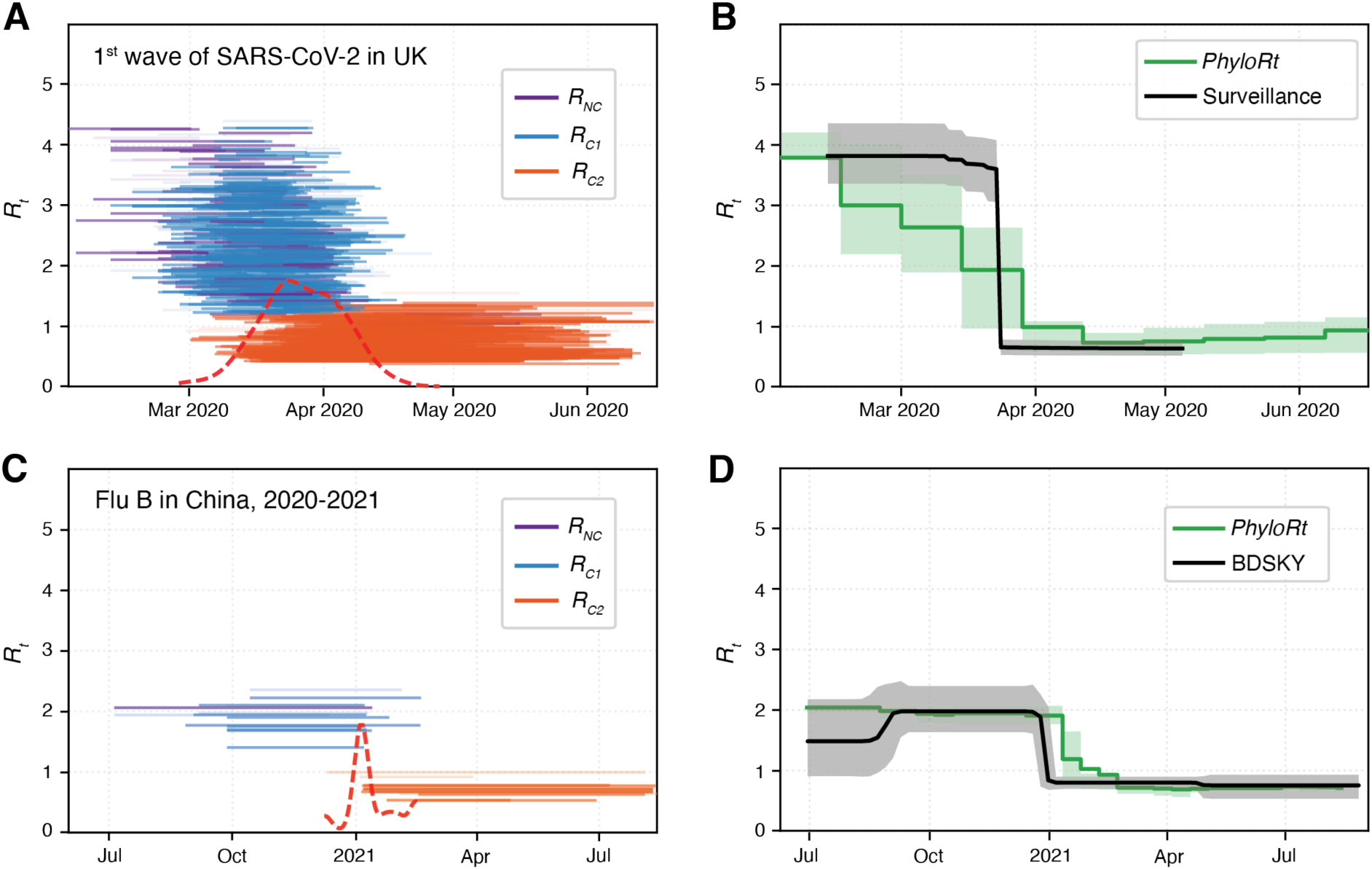
Application of *PhyloRt* to real outbreaks. **(A)** Subtree-level *R* inferred by *PhyloRt* using 50-tip subtrees of the first wave of SARS-CoV-2 in the UK, during early 2020. Each horizontal segment represents an estimate from an individual subtree, coloured by *R_NC_*, *R_C1_*, and *R_C2_***. (B)** *R_t_* inferred by *PhyloRt* from the subtree estimates in (A), shown with the IQR, and compared with a mathematical epidemiology estimate (27). **(C)** Subtree-level *R* inferred by *PhyloRt* using 100-tip subtrees of influenza B/Victoria clade 3a1, circulating in China from 2020 to 2021. **(D)** Comparison of transmission dynamics inferred for influenza B in China using *PhyloRt*, with the IQR derived from the subtree estimates in (C), compared with a Bayesian phylodynamic reconstruction (*R_e_* from BDSKY with 95% confidence interval). The dashed red curves at the bottom of panels C and D show a kernel-smoothed density of inferred change-point times across subtrees, normalized and scaled for visualization.

### Influenza B in China

Although global non-pharmaceutical interventions (NPIs) against COVID-19 during 2020–2021 led to a marked reduction in seasonal influenza worldwide (28, 29), China nevertheless experienced a resurgence of influenza B/Victoria circulation throughout 2021, accounting for the majority of globally detected influenza viruses during the 2020–2021 season. This occurred in a relatively closed transmission system, with low domestic restrictions following local elimination of SARS-CoV-2, and maintained by prolonged international border closures (25). We analyzed influenza B/Victoria clade 3a1, comprising 423 hemagglutinin (HA) gene sequences sampled between 7 July 2020 and 1 September 2021, which circulated in mainland China and was mainly responsible for outbreaks there from late 2020 to early 2021. *PhyloRt* estimates derived from time-scaled phylogenies dated using LSD2 (18) were highly consistent with effective reproductive number (*R_e_*) estimates inferred using BDSKY (30) in BEAST2, across all subtree sizes examined (50, 100, 200, and 400 tips) (Figs. 4C-D and SI Appendix, Fig. S16). Notably, the *PhyloRt* pipeline is computationally efficient, with tree reconstruction, dating, and inference steps each completing in under one minute. In contrast, Bayesian phylodynamic inference using BDSKY requires approximately one minute per million MCMC steps and typically 50–100 million steps to achieve adequate mixing (ESS > 200), resulting in runtimes on the order of hours. Analyses involving larger numbers of sequences or longer genomes (e.g., RSV or SARS-CoV-2) can require substantially longer runtimes, potentially extending to days or even weeks, depending on convergence requirements and analysis setup.

## Discussion

In this study, we demonstrate that time-varying transmission dynamics can be inferred reliably from outbreak phylogenies using a scalable, subtree-based deep-learning phylodynamic framework. Across extensive simulations and several empirical data sets from human respiratory pathogens, we show that accurate inference depends critically on aligning training conditions with realistic epidemiological and phylogenetic properties, including the birth–death model, the expected molecular evolutionary (phylogenetic) resolution of the data, and the tree-dating procedure (SI Appendix, Figs. S13 C-D and S.2) used to generate time-scale phylogenies. By decomposing large phylogenies into overlapping subtrees, *PhyloRt* captures temporal changes in transmission while remaining computationally efficient on ultra large datasets. Furthermore, applications to SARS-CoV-2 and influenza B show that *PhyloRt* can recover transmission dynamics from poorly resolved phylogenies that are consistent with estimates from mathematical epidemiology and current Bayesian phylodynamic methods.

Recent advances in deep-learning–based phylodynamics have demonstrated the potential to infer epidemiological parameters at unprecedented scales (14, 16). However, most existing studies rely on transmission trees simulated under birth–death models as training data, even though such trees cannot be reliably reconstructed from outbreak genomic data alone. Our previous work showed that incorporating partial contact tracing information can substantially refine tree topology and improve inference accuracy (17); however, in practice such data are rarely available, limiting their applicability. *PhyloRt* facilitates the application of such frameworks to real outbreak settings by operating directly on poorly resolved phylogenies generated by routine genomic surveillance.

Further, our results show that deep-learning–based phylodynamic inference is highly sensitive to the realism of training data. In particular, training under a basic birth–death model for pathogens that exhibit incubation periods or superspreading—features that characterize many respiratory viruses—leads to systematic underestimation of the reproductive number (Fig. 1B). This finding has implications beyond deep learning (e.g. for likelihood-based methods), suggesting that phylodynamic analyses based on overly simplified epidemiological assumptions may introduce bias when applied to real outbreak data (13, 31, 32).

Estimates of epidemic dynamics derived from pathogen genomic data, as an independent data source, can complement and augment traditional epidemiological inference from case surveillance, which can be limited by reporting biases or incomplete data that obscure true transmission trends (9). Although in our study misspecification of the sampling proportion can introduce directional bias in estimated reproductive numbers, we find that change-point detection remains robust (SI Appendix, Figs. S13 A-B and S.2), indicating that overall temporal transmission dynamics can be reliably reconstructed. In addition, the computational efficiency of *PhyloRt* permits inference to be evaluated quickly across multiple plausible sampling-proportion scenarios. We find that temporal transmission dynamics remain recoverable even under low sampling intensities, on the order of 0.1% of infections (SI Appendix, Figs. S14 and S.2), suggesting that routine sequencing data can be sufficient to reconstruct transmission dynamics (33, 34).

*PhyloRt* is designed to be applied to local transmission lineages, which are monophyletic clades derived from an importation event into the study population. As demonstrated in our analyses, local transmission lineages can be often identified and isolated from global phylogenies, which reduces the confounding effects of combining multiple local lineages into a single phylogeny (35). In addition, the subtree-based design of *PhyloRt* naturally supports incremental updating as new sequences are added, particularly when combined with tools such as UShER (36) that can efficiently place new genome sequences onto existing large trees. Future work will explore applying *PhyloRt* to track emerging variants with lineage-specific variation in transmission rates, and will assess the effects of progressively incorporating new sequence samples on inference robustness.

Our study has several limitations. First, the choice of subtree size involves an inherent trade-off between temporal resolution and estimation stability, and the optimal choice depends on both the size of the full phylogeny and the epidemiological context. Although our simulation results provide some practical guidance, real outbreaks may exhibit successive and step-wise changes in transmission dynamics (e.g. arising from introduction of non-pharmaceutical interventions (27)). In such settings, small subtrees (for example, 50 tips) can better capture rapid changes, even in large phylogenies of >10,000 sequences, as observed in the UK SARS-CoV-2 dataset we analysed (Figs. 5A-B and SI Appendix, Fig. S15). However, very small subtrees may lead to increased estimation uncertainty under certain conditions.

Second, our training framework assumes a single change-point per subtree. In practice, final inference relies on aggregation across subtrees within a piecewise-constant formulation, which is identifiable (24) and helps mitigate the impact of individual prediction errors, allowing multiple temporal segments to be approximated at the full-tree level. As in other phylodynamic methods, closely spaced changes in *R_t_* may not be well resolved; the subtree-based formulation further limits temporal resolution. Similarly, sampling proportion is assumed constant within each subtree, and time-varying sampling is captured only at the complete-tree level through aggregation. Extending our framework to accommodate multiple change points in training and inference remains challenging and warrants further investigation.

Third, *PhyloRt* focuses on estimating the reproductive number, *R_t_*, while other epidemiological parameters, such as infectious/incubation period or superspreading parameters, are not explicitly inferred. These parameters are difficult to estimate reliably from poorly resolved phylogenies (SI Appendix, Figs. S1-2 and S.1) (17), limiting the scope of inference. Finally, we did not benchmark against BEAST/BEAST2 on the simulated fixed trees, as in previous studies (14, 16, 17, 37), because *PhyloRt* operates on pathogen-specific realistic trees dated using LSD2 (18), for which Bayesian phylodynamic methods are not well suited (17).

Overall, these findings highlight the potential of scalable deep-learning–based phylodynamic frameworks, trained on pathogen-specific realistic phylogenies, to complement existing epidemiological and phylogenetic approaches for outbreak analysis. As genomic surveillance continues to expand, such frameworks may play an increasingly important role in interpreting pathogen genomic data at scale.

## Materials and Methods

### Simulations for neural network model training and testing

We simulated transmission trees under two epidemiological scenarios used for deep learning model training and testing: constant reproduction number in which *R_t_* remains constant through time, and time-varying reproduction number, in which *R_t_* shifts abruptly at one change point. All simulations were performed using *treesimulator* (v0.2.20; (38)), and epidemiological parameter ranges are summarized in SI Appendix, Table S1.

For constant transmission rates, trees were generated under four BD model variants: BD, BDEI, BDSS, and BDEISS, using a multi-type birth–death (MTBD) approach (Fig. 1B). For each model, we simulated 100,000 trees for training and 2,000 trees for testing, with tip counts uniformly sampled between 200 and 500. Simulated time-scaled transmission trees were converted into genetic-distance trees by assigning branch lengths *B* using a Poisson process,

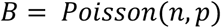

where *n* denotes the sequence length and *p* is the product of the evolutionary rate (in substitutions per site per year) and the time-scaled branch length. Zero-length branches were collapsed and resulting polytomies were randomly resolved by iteratively pairing descendant lineages to generate a fully bifurcating tree, following a coalescent-like random branching process. Genetic-distance trees were subsequently re-dated using LSD2 (18), with a minimal branch length of 1 hour, to obtain realistic, pathogen-specific phylogenies with limited resolution. Tip dates were obtained from the time-scaled transmission trees; to avoid unrealistically early root ages, the root time was constrained to lie within ±1 day of the reference time. The molecular clock rate used during re-dating was fixed to the same value applied in the genetic-distance transformation.

To compare the impact of different birth–death model formulations, simulations were first parameterized using SARS-CoV-2, producing trees consistent with its genome length and evolutionary rate (SI Appendix, Table S2) under each BD variant. To examine pathogen-specific effects, we additionally generated pathogen-specific trees for seasonal human influenza (A/H3N2, A/H1N1pdm09, influenza B) and respiratory syncytial virus (RSV A and B). For these analyses, 100,000 training trees and 2,000 testing trees were simulated under a birth–death model matched to each pathogen’s transmission dynamics (BDEI for influenza and RSV) and transformed using the corresponding genome lengths and evolutionary rates (SI Appendix, Table S2).

For time-varying transmission rates, we simulated 10,000 large trees under BDEI and BDEISS, with total tip counts ranging from 1,000 to 10,000. The change point was determined from simulations under constant *R*, defining the tree height; a second simulation was then performed from time zero with a single change point introduced at this time. At the change point, *R* (and superspreading parameters for BDEISS) were resampled from predefined epidemiological parameter ranges (SI Appendix, Table S1), while all other epidemiological parameters were held constant including sampling proportion. The resulting trees were then transformed into pathogen-specific (H3N2, influenza B, and SARS-CoV-2) phylogenies as described above.

Subtrees were extracted by pruning descendant tips from internal nodes of the large trees. Only internal nodes with at least 21 tips were considered. For target sizes n = 50, 100, 200, and 400, subtrees were generated as follows: if an internal node had at least n descendants, the first n descendants were retained; if it had fewer than n but more than the previous lower target size, all descendants were retained and assigned to the n-tip category. Consequently, subtrees in the 50-, 100-, 200-, and 400-tip categories comprised 21–50, 51–100, 101–200, and 201–400 tips, respectively.

For each subtree size group, we selected one root subtree, and one randomly chosen recent subtree without a change point, together with 20 randomly sampled subtrees with and without a change point separately. Subtrees containing a change point were required to have the change point located between 20% and 80% of the subtree’s temporal span. This procedure yielded, per subtree size group, 100,000 training subtrees and 2,000 testing subtrees with a change point, and the same numbers without a change point.

### Feature representation

We used the PhyloCNN framework (16) to represent phylogenetic trees and to train neural network models for phylodynamic inference. All time-scaled trees were first rescaled such that the mean branch length equalled 1 (14). Each tree was then encoded into two feature tables corresponding to internal nodes and leaf nodes, which were concatenated to form a matrix of dimension 2n × 19, where n denotes the number of tips in the tree. The encoding follows a 2-neighborhood design: node features incorporate information from the node’s great-grandparents (parents of grandparents) and its grandchildren, together with the sampling proportion of the tree. If a tree contained fewer than n tips, zero padding was applied to reach the target size.

For subtrees extracted from large trees with a single change point, we additionally added two tree-level temporal features that encode the subtree’s relative position within the full tree and capture potential conditional sampling effects (known in the birth-death literature of “pull-of-the-present” (39)):

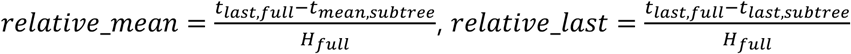

where 𝑡_last, full_ is the most recent (last) sampling date in the full tree, 𝑡_mean, subtree_ and 𝑡_last, subtree_ are the subtree mean and last sampling dates, respectively, and 𝐻_full_ is the full tree height. These features were appended as two additional columns with constant values across all rows, analogous to the sampling-proportion feature, yielding a 2n × 21 representation. Incorporation of these temporal features resulted in consistent improvements in neural network performance across both classification and regression tasks (SI Appendix, Fig. S17).

### Neural network models training and evaluation

Neural network models were implemented in Python 3.8 using TensorFlow 1.5.0, Keras 2.2.4, and scikit-learn 0.19.1. Models were trained on PhyloCNN encodings with input tensors of dimension (n × 19 × 2) or (n × 21 × 2), where n denotes the predefined tree size and the two channels correspond to internal-node and leaf-node representations. The neural network architecture comprised a three-layer convolutional feature extractor followed by a fully connected prediction head. The convolutional module consisted of one grouped convolutional layer (separate kernels for internal-node and leaf-node channels) with 32 filters, followed by two pointwise (1×1) convolutional layers with 32 filters each. All convolutional layers used ELU activation and were interleaved with batch normalization, followed by global average pooling. The pooled features were passed to four fully connected hidden layers arranged in a funnel shape with 64, 32, 16, and 8 neurons, respectively, all using ELU activation. The final output layer used linear activation for regression tasks and sigmoid activation for classification tasks.

Models were trained using the Adam optimizer. Mean absolute percentage error was used as the loss function for regression tasks, and binary cross-entropy for classification tasks. Training was conducted for up to 1,000 epochs with batch sizes of 512 (regression) or 1024 (classification), using a validation split of 0.3. Early stopping was applied based on validation loss with a patience of 100 epochs, and the best-performing weights were retained.

To assess birth–death model and pathogen-specific effects, regression models were trained on 100,000 simulated trees and evaluated on 2,000 independent trees under constant transmission rates for each pathogen and each birth–death model variant, with tip sizes ranging from 200 to 500. For subsequent *R_t_* estimation, models were trained on subtrees. For each subtree size group, a classification model was trained to distinguish trees with and without a change point using balanced datasets of 200,000 subtrees. Separate regression models were then trained to estimate epidemiological parameters, including the reproductive number for trees without a change point (*R_NC_*), and for trees with a single change point the reproductive number before (*R_C1_*) and after (*R_C2_*) the change point, as well as the change-point position within the subtree. Each regression model was trained using 100,000 subtrees corresponding to its respective task.

Regression performance was evaluated using the mean relative error (MRE) metric:

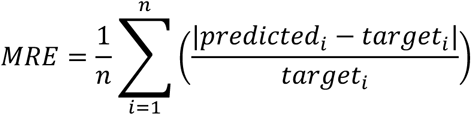

and the mean relative bias (MRB):

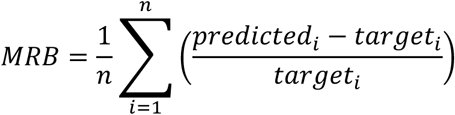

where n is the number of simulated trees used as the testing dataset. Classification performance was assessed using accuracy (ACC), sensitivity (SN), specificity (SP), precision (PPV), and negative predictive value (NPV) defined in terms of true positives (TP), true negatives (TN), false positives (FP), and false negatives (FN) as follows:

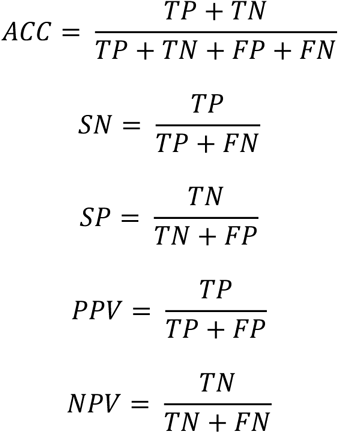

### Estimation of *R_t_* using *PhyloRt*

*PhyloRt* estimates time-varying transmission rates by aggregating predictions from multiple overlapping subtrees extracted from a single phylogenetic tree. Given an input tree, subtrees are first extracted from internal nodes using predefined target subtree (clade) sizes, as described in the simulation section above.

We adopt a hierarchical inference strategy (Fig. 2A). For each subtree, a classification model is first applied to assess the presence of a change point. If no change point is detected (classification probability < 0.1), a regression model is used to estimate a single reproductive number *R_NC_*. If a change point is detected (classification probability > 0.9), separate regression models are used to estimate the reproductive number before (*R_C1_*) and after (*R_C2_*) the change point, together with the relative position of the change point within the subtree. Subtrees with intermediate classification probabilities (0.1–0.9) are excluded; thresholds can be relaxed if too few subtrees are retained.

*PhyloRt* then constructs a piecewise-constant estimate of *R_t_* by aggregating subtree-level predictions across time. The time span of the input tree is discretized into consecutive time bins of fixed width (14 days in this study; see SI Appendix, Fig. S18; smaller bins are noisier and larger bins smoother). Each subtree contributes to all time bins that overlap its temporal interval. For subtrees without a change point, *R_NC_* is assigned to all overlapping bins. For subtrees with a change point, the change-point time is compared with the midpoint of each bin, such that bins before the change point use *R_C1_* and bins after the change point use *R_C2_*.

Each subtree–bin contribution was assigned an effective weight:

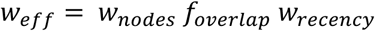

where 𝑤_nodes_ is the node-based weight, defined as the proportion of internal nodes and tips in the subtree relative to the total within the corresponding time span, and 𝑓_overlap_ is the fraction of the time bin covered by the subtree. The recency-based weight is optional and can be used to reduce the influence of time-averaged contributions when short-term dynamics are of interest. When applied,

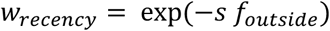

where 𝑓_outside_ denotes the fraction of the subtree’s temporal span lying outside the time bin and s > 0 controls the strength of recency weighting, with larger values placing greater emphasis on short-term contributions.

For each time bin, weighted contributions from all overlapping subtrees are aggregated to produce a point estimate of *R_t_* as a weighted mean, with the interquartile range (IQR; 25th–75th percentiles) derived from the corresponding weighted empirical distribution.

### Simulations for evaluation under time-varying transmission

We conducted additional simulations under four time-varying transmission scenarios, namely constant *R_t_*, increase in *R_t_*, decrease in *R_t_*, and alternating *R_t_* values, which were used exclusively to evaluate *R_t_* estimation. All scenarios assumed an incubation period of 2 days and an infectious period of 7 days. In the constant scenario, the effective reproductive number remained fixed at *R* = 1.5. In the increase scenario, *R* increased from 1.5 to 3 at day 120, whereas in the decrease scenario it declined from 2.5 to 1 at day 60. In the zigzag scenario, *R* alternated sequentially between 3, 0.5, 1.5, and 0.75 at days 45, 75, and 105. Simulations were performed under both BDEI and BDEISS models. For BDEISS, superspreading parameters were fixed at *X_ss_* = 15 and *f_ss_* = 0.1. Parameter values were chosen to span epidemiological ranges relevant to the pathogens considered in this study (23, 40). Transmission trees were generated with 2,000 to 3,000 tips (sampling proportion = 0.1) and 5,000 to 10,000 tips (sampling proportion = 0.3), with simulation end times ranging from 150 to 180 days. Each configuration was replicated independently three times, and resulting trees were transformed into realistic phylogenies parameterized for H3N2 influenza A, influenza B, and SARS-CoV-2.

Performance of *R_t_* estimation was assessed using time-normalized mean absolute error (MAE), computed with time weighting. Let *R̂*(𝑡) and 𝑅(𝑡) denote the estimated and true transmission rates at time *t* and let Δ𝑡 denote the width of each time bin (14 days). The MAE was defined as the integrated absolute error normalized by the total time span,

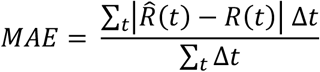

### Application to SARS-CoV-2 outbreak data from the United Kingdom in early 2020

We used genomic and epidemiological data from the first wave of SARS-CoV-2 infection in the United Kingdom during early 2020, which was unusually well represented by viral genomic sampling. Rapid fluctuations in viral importation during this period resulted in the emergence of more than 1,000 distinct local transmission lineages. To focus on sustained local transmission dynamics, we selected the 50 largest UK transmission lineages identified previously (6). These lineages were sampled between 2 February and 17 June 2020 and ranged in size from 86 to 1,865 tips.

For each lineage, a monophyletic maximum likelihood (ML) tree was pruned from the large tree (6) using Gotree (41). Branches corresponding to fewer than one mutation were collapsed, and any resulting polytomies were randomly resolved to produce binary trees. The resulting phylogenies were dated using LSD2 (18), applying the same SARS-CoV-2 molecular clock assumptions (Fig. 1C) and enforcing a minimum branch length of 1 hour.

Sampling proportions were obtained from our prior study (6) as weekly time-varying estimates, computed as the ratio of sequenced genomes to weekly estimated UK infections (27). When pruned subtrees spanned multiple sampling-proportion intervals, sampling proportions were assigned at the tip level and then averaged across tips within each subtree for use in *PhyloRt* inference. In total, 13,883 SARS-CoV-2 genome sequences from the selected lineages were included in the analysis. To contextualize the estimates of *R_t_* inferred from genomic data by *PhyloRt*, we compared them with estimates derived from a mathematical epidemiological model applied to time series incidence data in the UK from the same study period (27).

### Application to flu B circulation in China in 2020-2021

We also applied *PhyloRt* to influenza B virus circulation in China during 2020–2021 and compared the *PhyloRt* estimate to that obtained using the phylodynamic inference framework implemented in BEAST2 (26). During this period, China maintained a strict COVID-19 elimination strategy while experiencing a resurgence of seasonal influenza activity, with no evidence of international introductions of human seasonal influenza, resulting in a relatively closed transmission system (25). We analyzed 423 hemagglutinin (HA) gene sequences from influenza B lineage B/Victoria clade 3a1, sampled between 7 July 2020 and 1 September 2021. Maximum-likelihood phylogenies were reconstructed using IQ-TREE (v.2) (42) and processed as described above for the SARS-CoV-2 analysis, including branch collapsing, random resolution of polytomies, and time scaling using LSD2 (18) under the same influenza B molecular clock assumptions (Fig. 1C), with a minimum branch length of 1 hour. Bayesian phylodynamic estimates were taken from our prior study (25) using BEAST2 (26) with the birth–death serial skyline (BDSKY) model (30) to infer the effective reproductive number (*R_e_*). The constant sampling proportion used in *PhyloRt* inference was obtained from the BDSKY analysis.

## Data Availability

All data produced are available online at https://github.com/xieruopeng/PhyloRt

## Acknowledgments

The computations were performed using research computing facilities offered by Information Technology Services, the University of Hong Kong. We thank Dr. Louis du Plessis for his valuable comments and fruitful discussions on the manuscript. This work was supported by the Marie Skłodowska-Curie Actions (Project No. 101203810) and the Research Grants Council of the Hong Kong Special Administrative Region, China (Project No. HKU PDFS2425-7S01) (R.X.). O.G.P. and M.U.G.K. are supported by the Oxford Martin School Programme for Pandemic Genomics. M.U.G.K. acknowledges additional support from The Rockefeller Foundation (PC-2022-POP-005); the Health AI Programme from Google.org; the Oxford Martin School Programme in Digital Pandemic Preparedness; the European Union’s Horizon Europe programme (MOOD, Grant No. 874850; E4Warning, Grant No. 101086640); the Wellcome Trust (Grants 303666/Z/23/Z, 226052/Z/22/Z, and 228186/Z/23/Z); UK Research and Innovation (Grant APP8583); the Medical Research Foundation (Grant MRF-RG-ICCH-2022-100069); UK International Development (Grant 301542-403); the Bill & Melinda Gates Foundation (Grants INV-063472 and INV-090281); and the Novo Nordisk Foundation (Grant NNF24OC0094346). The funding bodies had no role in the design of the study; the collection, analysis, or interpretation of data; or the writing of the manuscript. The contents of this publication are the sole responsibility of the authors and do not necessarily reflect the views of the European Commission or other funders.

## Data and materials availability

All anonymized data, code, and analysis files are available in the GitHub repository (https://github.com/xieruopeng/PhyloRt).

