## Supplementary materials for "Scalable deep-learning-based inference of time-varying transmission dynamics from outbreak phylogenies"

**This file includes:**

Supporting Information Text
Figs. S1 to S18
Tables S1 to S6
SI Reference

### Supporting Information Text

**S.1 Impact of birth–death model specification on neural network performance.** Using SARS-CoV-2 as an example, we generated trees under BD, the birth–death model with exposed and infectious classes (BDEI), the birth–death model with superspreading (BDSS), and the birth–death model with both exposed–infectious structure and superspreading (BDEISS), the latter most closely reflecting SARS-CoV-2 transmission dynamics (Fig. 1B). Neural networks were trained separately on realistic trees characterised by SARS-CoV-2 sequence length and evolution rates under each BD variant, with a number of mutations per year as shown in Fig. 1C and evaluated across all combinations (Fig. 1B and SI Appendix, Figs. S1-2). Substantial discrepancies emerged when models were applied to a different tree type than the one used for training. Models trained on simpler trees (e.g., BD) showed higher mean relative errors (MRE) when tested on more complex models (e.g., BDEI, BDSS, BDEISS) and tended to underestimate $R_0$, resulting in a negative mean relative bias (MRB). The best performance, with near-zero MRB, was consistently observed when training and testing used the same tree type. These patterns were also confirmed using transmission trees by both maximum likelihood methods and deep-learning approaches, the latter based on summary-statistic features, across a broader range of birth–death models. Superspreading parameters remained difficult to estimate (1), and incorporating incubation periods (as in BDEI and BDEISS) further reduced accuracy for both infectious and incubation period estimates (MRE > 0.2), reflecting the limits of low phylogenetic resolution (SI Appendix, Fig. S1).

**S.2 Sensitivity of neural network predictions to sampling proportion and tree-dating methods.** Neural network models take as input a sampling proportion together with a time-scaled phylogenetic tree, and we therefore evaluated the sensitivity to variation in both inputs. Here, we used models trained on 200-tip subtrees pruned from 10,000 large trees (1,000 to 10,000 tips) simulated under the SARS-CoV-2 parameterization, with a testing dataset of 4,000 trees, comprising 2,000 trees with a change point and 2,000 trees without a change point. This analysis was limited to individual subtree inputs to the neural network models and did not involve the full hierarchical *PhyloRt* inference.

To evaluate sensitivity to sampling proportion, we perturbed the sampling proportion of the testing trees by -50%, -25%, +25% and +50% relative to the true value. The classification model was evaluated against the true change-point labels, whereas regression sensitivity was quantified by relative differences from predictions obtained using the true sampling proportion. As the sampling proportion deviated from its true value (SI Appendix, Figs. S13 A-B), classification performance and estimation of the change-point position remained largely stable. In contrast, sampling proportion introduced a directional bias in regression outputs for $R_{NC}$, $R_{C1}$, and $R_{C2}$, with $R$ estimates decreasing with lower sampling proportions and increasing with higher sampling proportions. These results indicate that sampling proportion primarily affects the absolute scale of inferred $R$ values, rather than the temporal dynamics recovered by *PhyloRt*.

In addition, we assessed sensitivity to the underlying sampling level across different true sampling proportions. Because the training dataset was designed to span the widest feasible range of sampling proportions, we extended the lower bound to 0.001 (0.1%) (SI Appendix, Table S1), the lowest sampling proportion at which birth–death simulations can reliably generate phylogenetic trees of fixed tip size. Model performance was also evaluated across this range using sampling-proportion intervals of width 0.2 for the classification model and intervals of width 0.1 for the regression models. Despite this scaling effect, overall model performance remained stable across a wide range of sampling proportions (SI Appendix, Fig. S14), including values as low as 0.1%, suggesting that highly incomplete sampling can still support reconstruction of transmission dynamics.

To assess sensitivity to the tree-dating method, we compared predictions from trees dated using LSD2 (2), TreeTime (3), and Chronumental (4), with LSD2 used as the default for model training and baseline testing (SI Appendix, Figs. S13 C-D). For consistency, all methods used the same SARS-CoV-2 genome length and clock rate; a clock-rate standard deviation was specified for LSD2 and TreeTime but not for Chronumental. Stochastic resolution of polytomies was applied in TreeTime, and 200 optimization steps were used for Chronumental. Any remaining unresolved polytomies in trees dated using TreeTime or Chronumental were randomly resolved prior to input into the neural network models.

Relative to LSD2, predictions obtained from trees dated using TreeTime or Chronumental showed consistently reduced performance, with lower PPV and NPV for classification and higher relative errors for regression. Error distributions were also broader, indicating increased variability. Together, these results suggest that differences in how tree-dating methods resolve branch lengths and polytomies can introduce systematic mismatches between training and inference conditions, underscoring the importance of using consistent tree-dating procedures for model training and application.

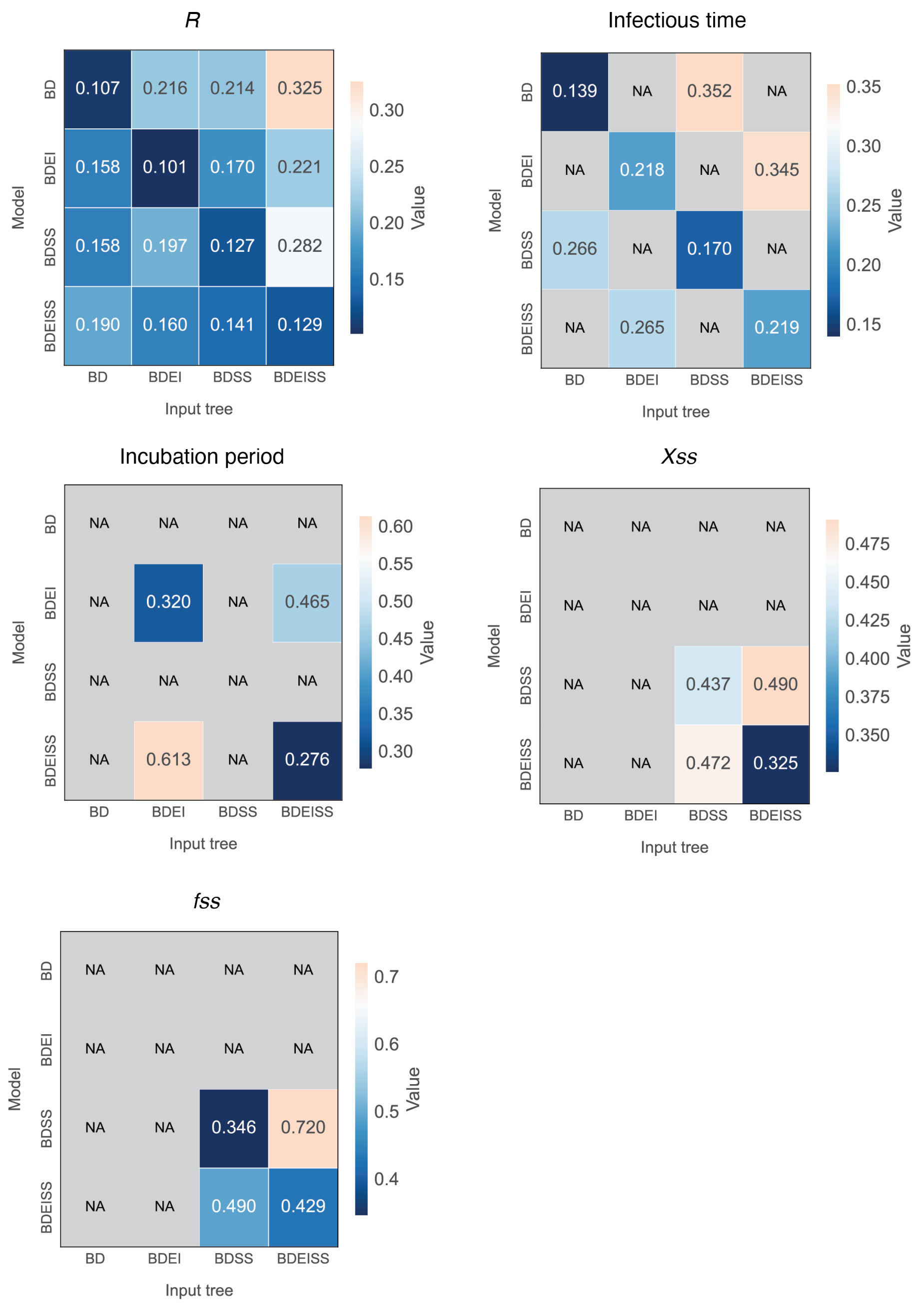


**Fig. S1.** Mean relative error (MRE) of parameter estimates. Models trained and tested on all combinations of poorly resolved trees characterised by SARS-CoV-2 sequence length and evolution rate under each BD variant. NA values indicate either parameter combinations for which estimates are not interpretable under the corresponding model assumptions or cases where no valid output was produced.

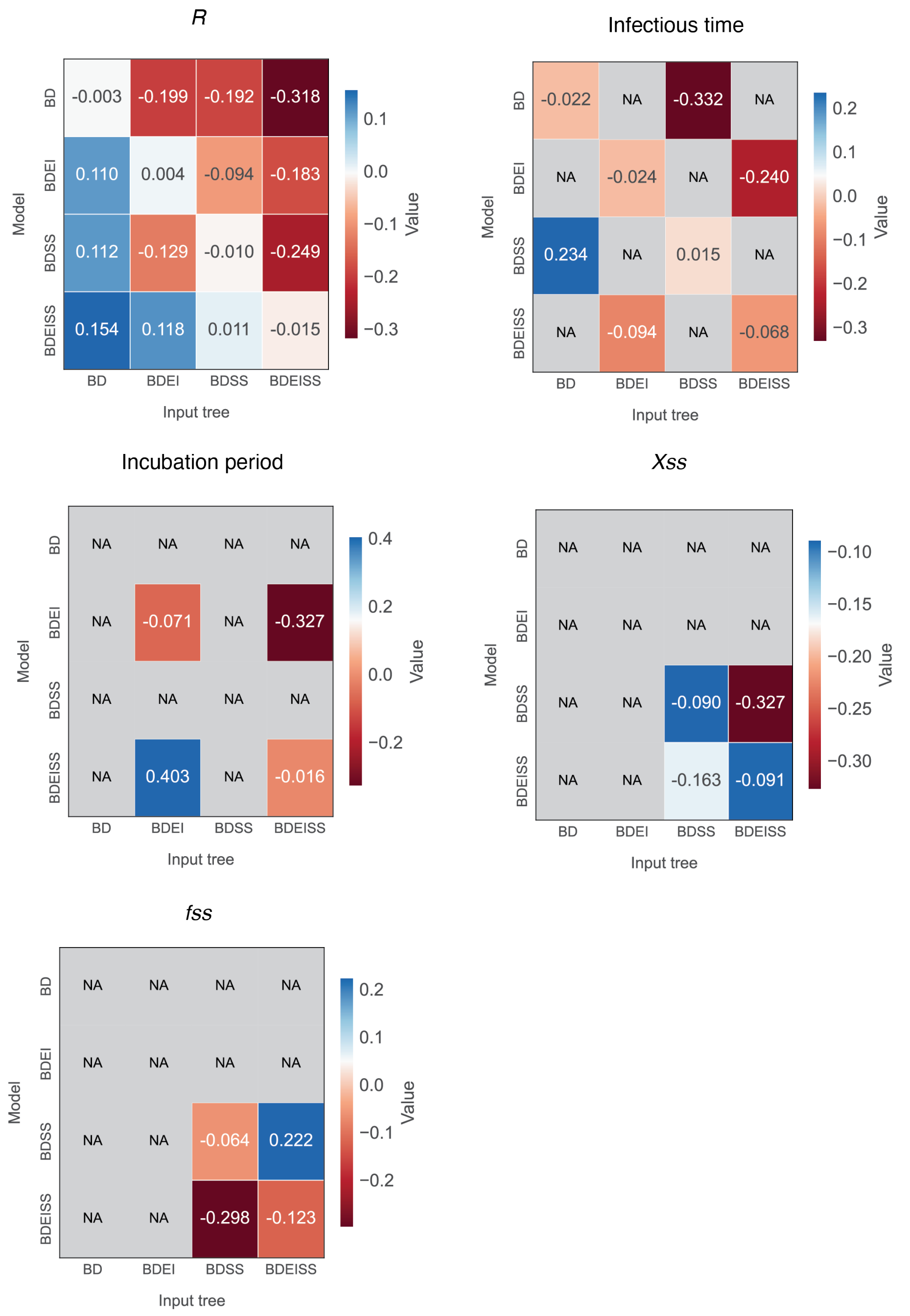


**Fig. S2.** Mean relative bias (MRB) of parameter estimates. Models trained and tested on all combinations of poorly resolved trees characterised by SARS-CoV-2 sequence length and evolution rate under each BD variant. NA values indicate either parameter combinations for which estimates are not interpretable under the corresponding model assumptions or cases where no valid output was produced.

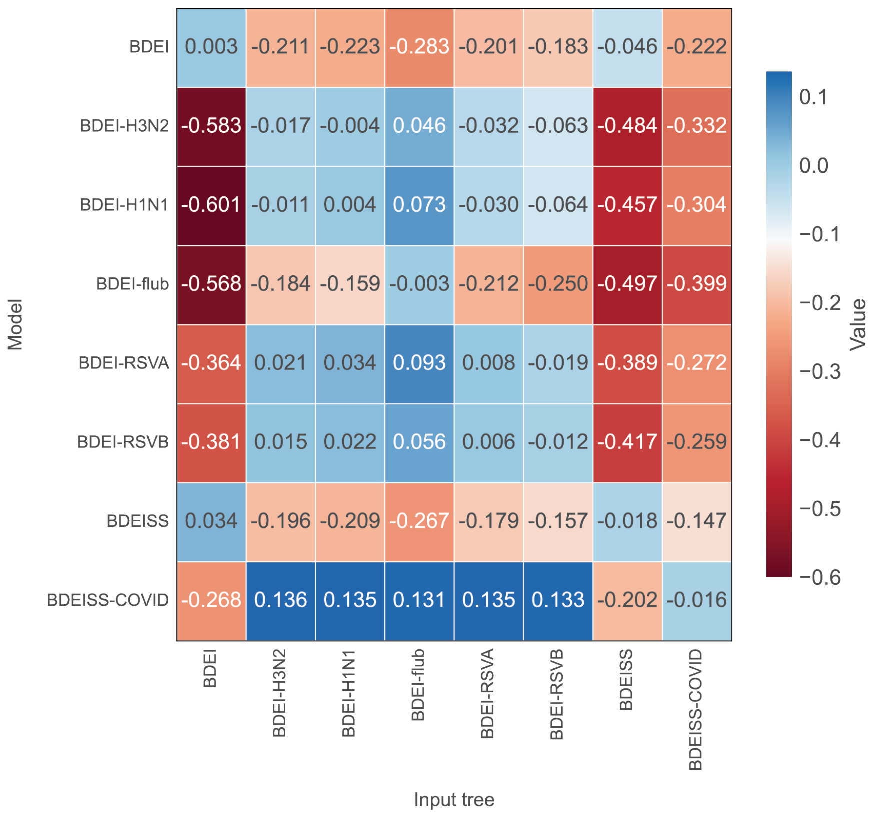


**Fig. S3.** Mean relative bias (MRB) of *R* estimates for pathogen-specific models trained on poorly resolved trees under biologically realistic birth–death models for each pathogen, evaluated across all model–input tree combinations.

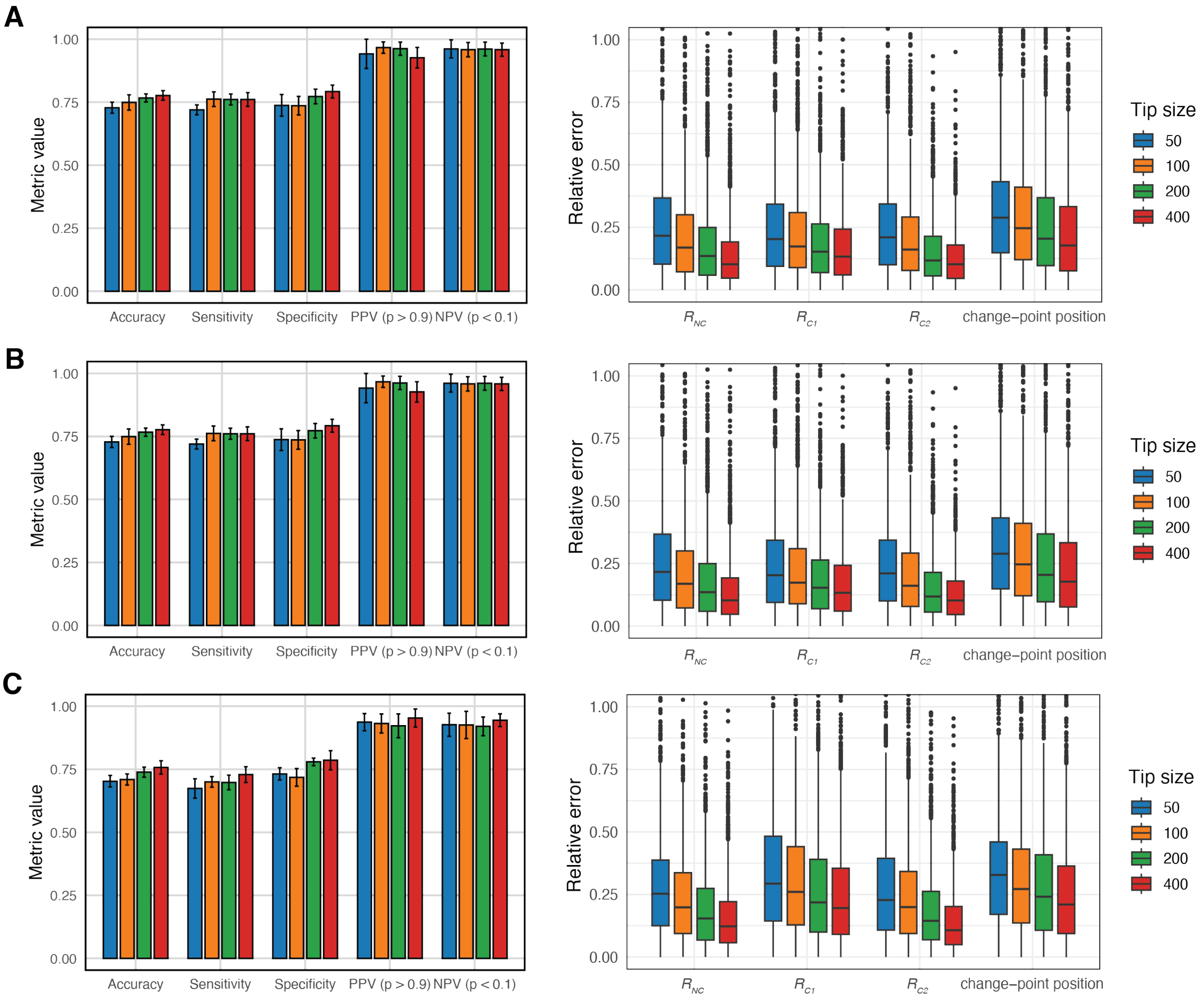


**Fig. S4.** Model performance of hierarchical subtree-based inference across subtree sizes. (A–C) H3N2, flu B, and SARS-CoV-2. Left panels show classification performance metrics stratified by subtree tip size. For accuracy (ACC), sensitivity (SN), and specificity (SP), predicted change-point probabilities were classified using a threshold of 0.5. Right panels show distributions of relative error for estimated $R_{NC}$, $R_{C1}$, $R_{C2}$, and change-point position across subtree sizes.

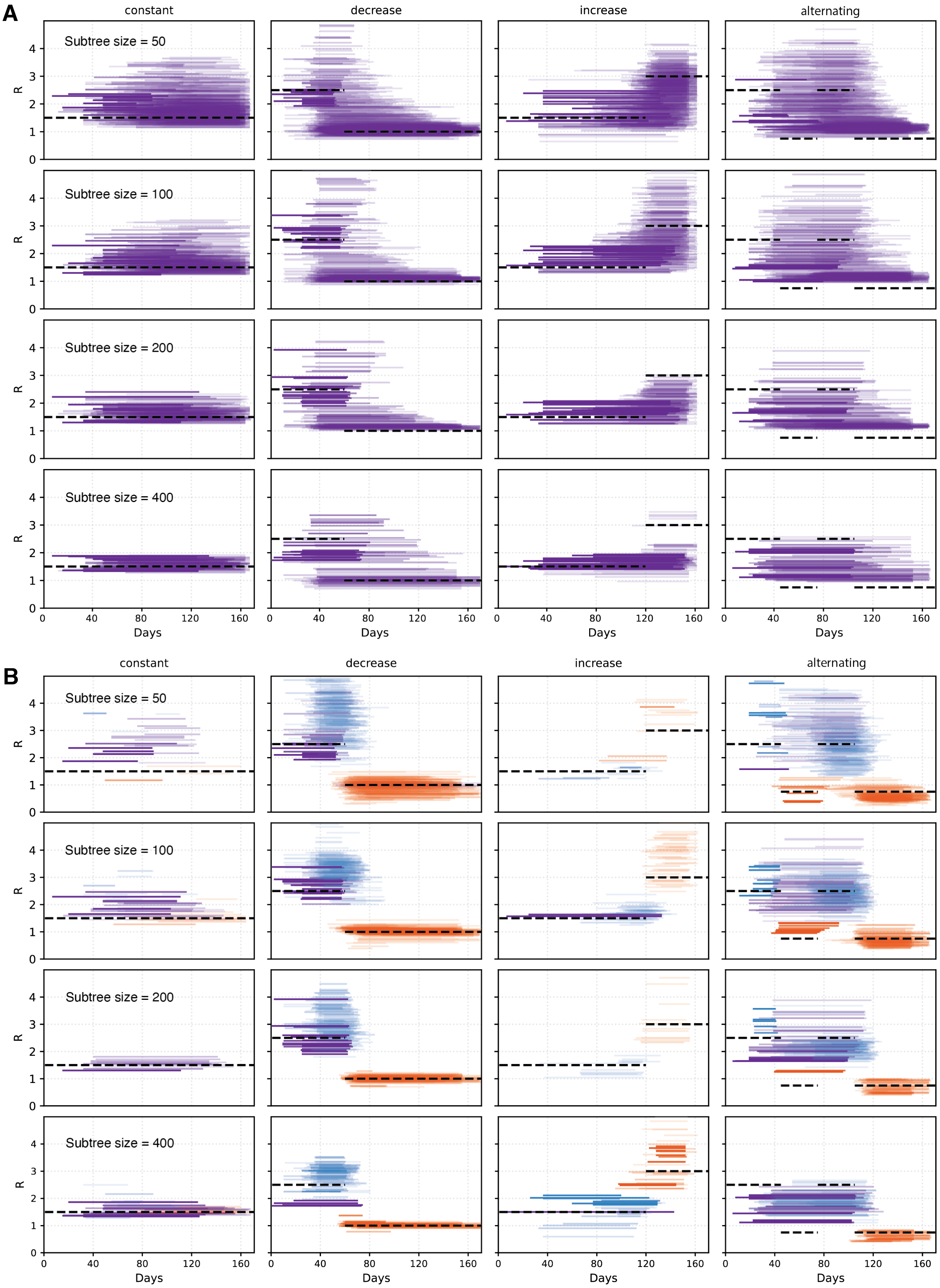


**Fig. S5.** Subtree-level reproductive number estimates for H3N2 simulated trees with 2,000–3,000 tips, using a baseline direct-averaging approach (A) and hierarchical inference (B) across transmission scenarios and subtree sizes.

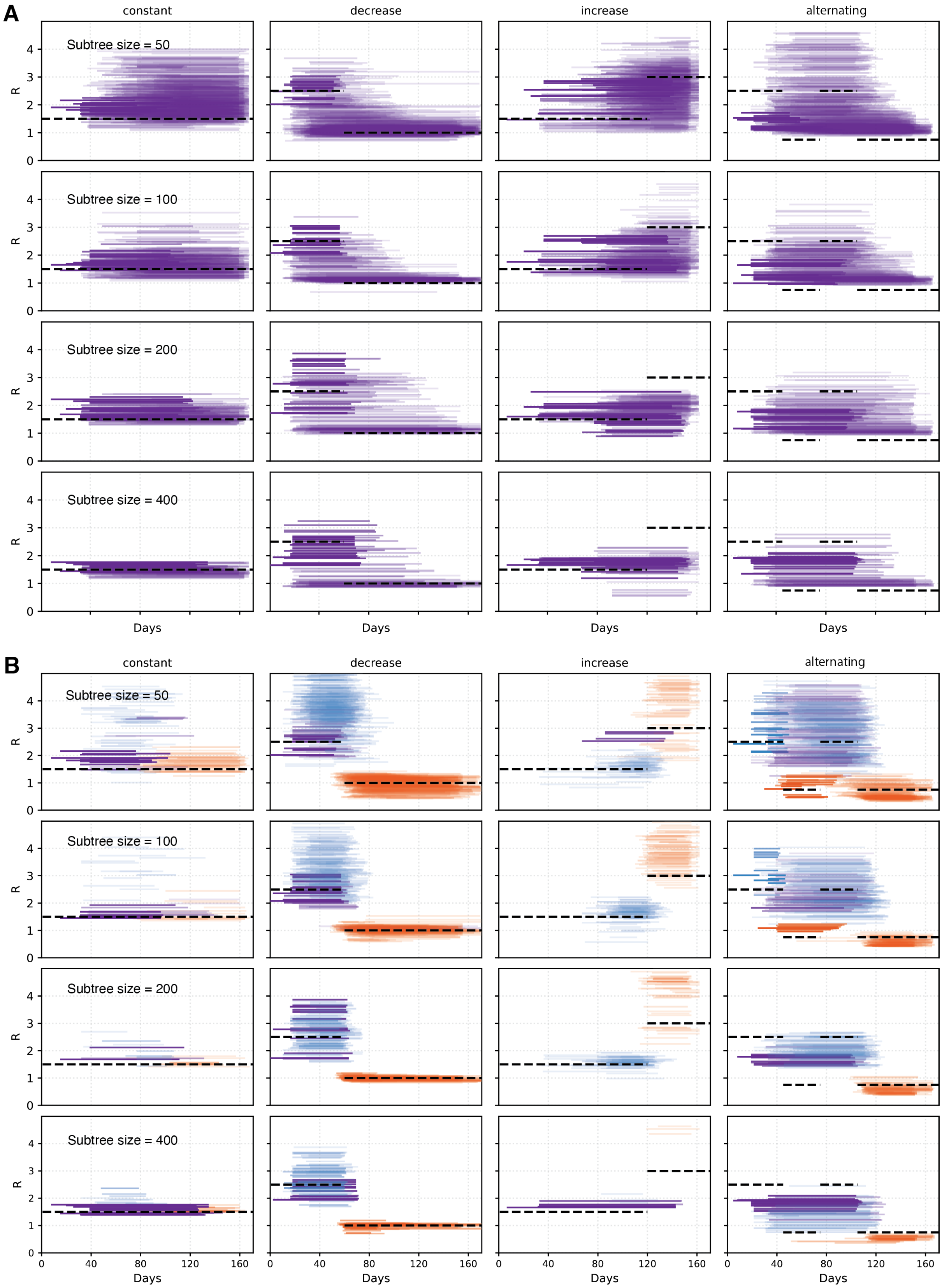


Fig. S6. Subtree-level reproductive number estimates for flu B simulated trees with 2,000–3,000 tips, using a baseline direct-averaging approach (A) and hierarchical inference (B) across transmission scenarios and subtree sizes.

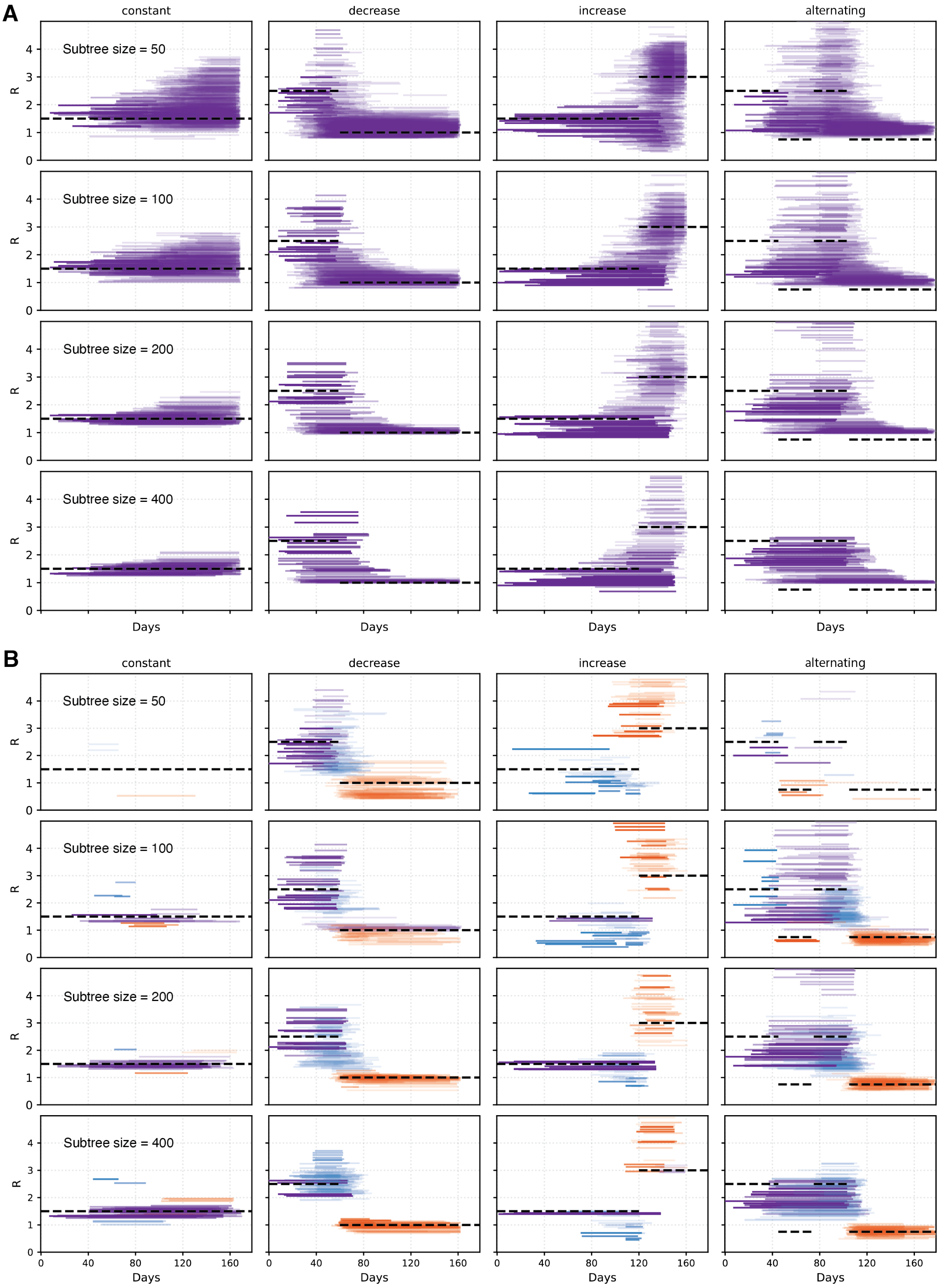


Fig. S7. Subtree-level reproductive number estimates for SARS-CoV-2 simulated trees with 2,000–3,000 tips, using a baseline direct-averaging approach (A) and hierarchical inference (B) across transmission scenarios and subtree sizes.

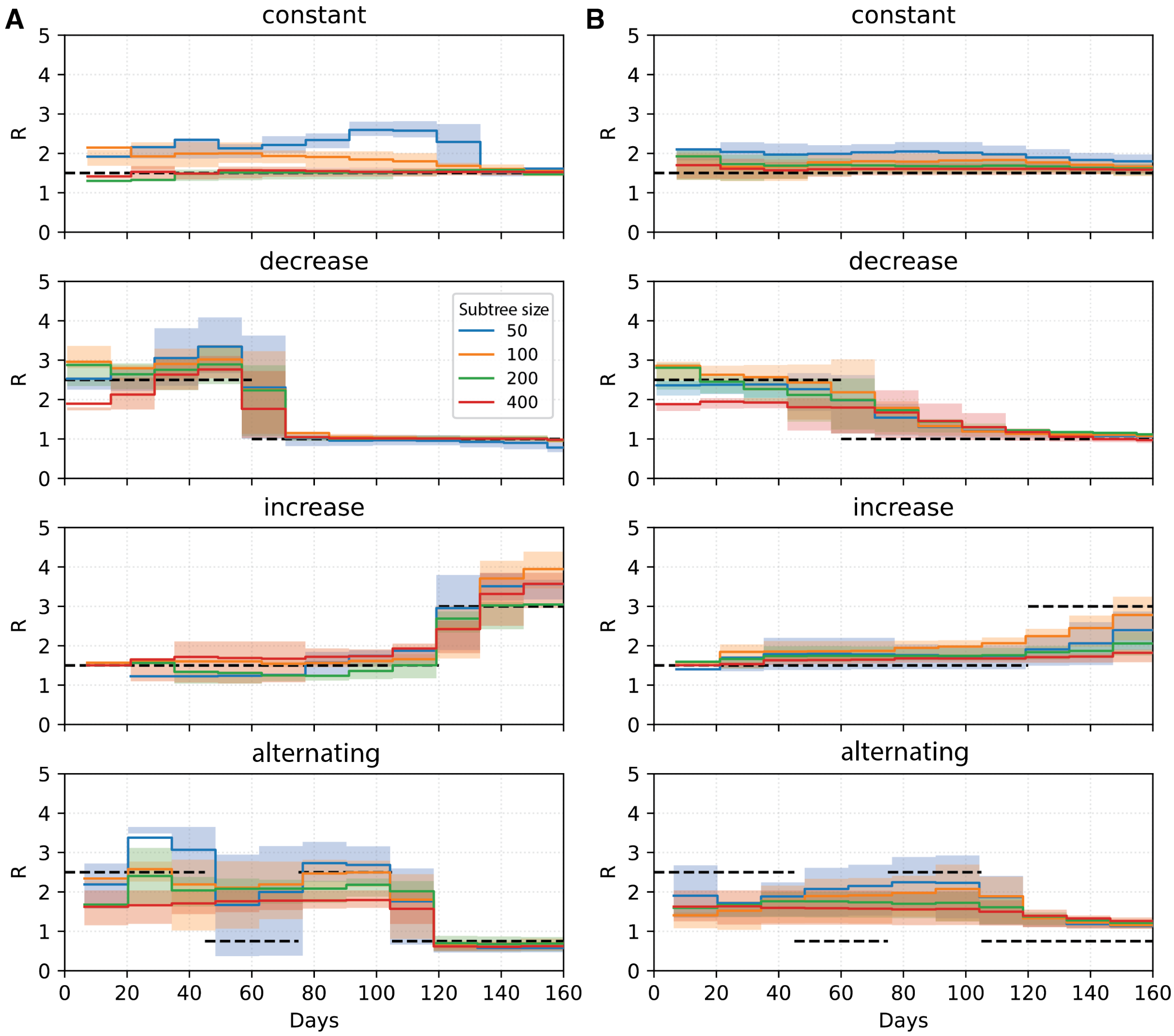


**Fig. S8.** Comparison of hierarchical (A) and baseline direct-averaging (B) subtree-based inference for estimating $R_t$ in three independently simulated H3N2 trees with 2,000–3,000 tips across four transmission scenarios. Colors indicate subtree size. Solid lines show the estimated mean $R_t$, with shaded bands representing the IQR. Dashed black lines denote the true underlying reproductive number.

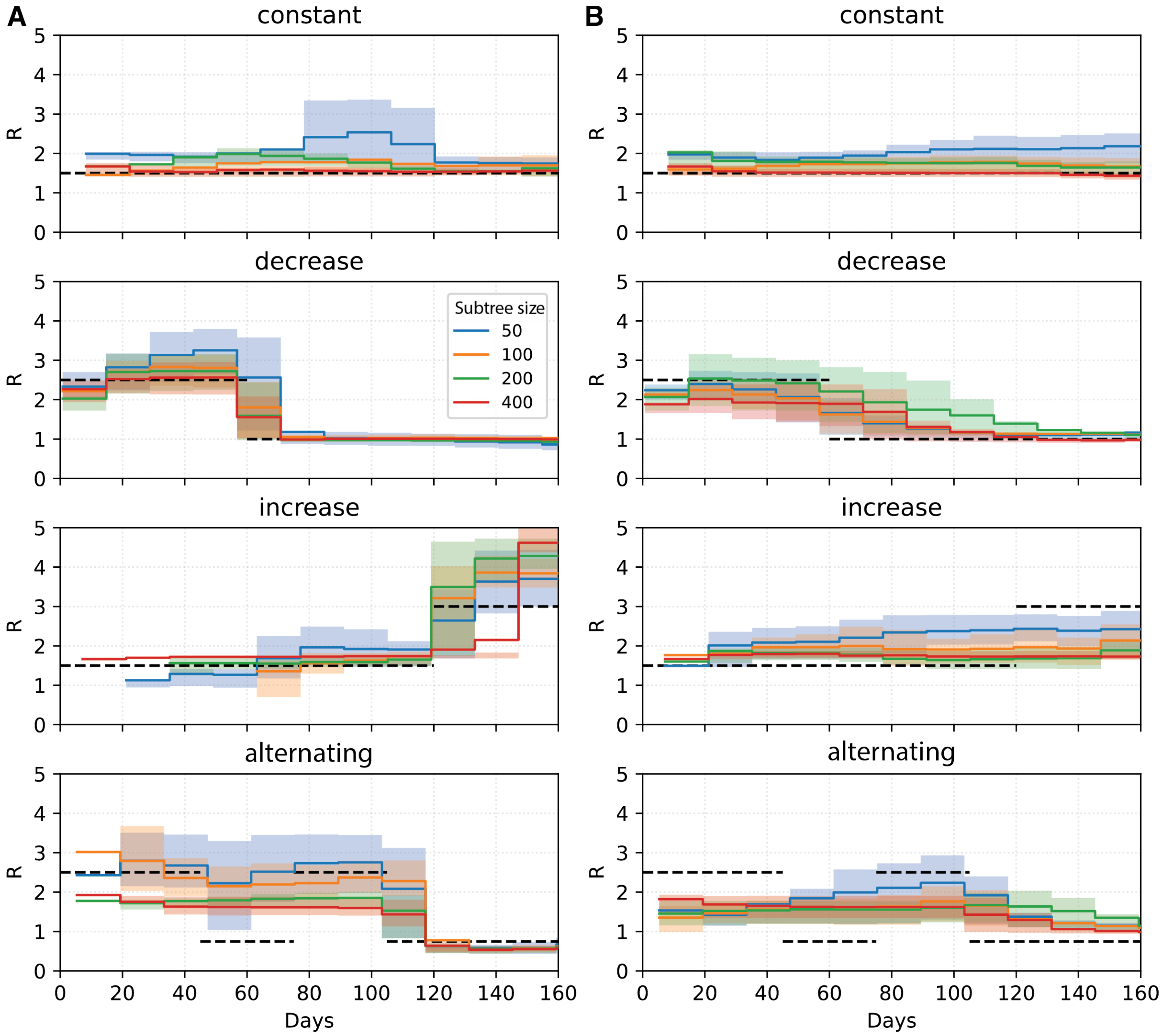


**Fig. S9.** Comparison of hierarchical (A) and baseline direct-averaging (B) subtree-based inference for estimating $R_t$ in three independently simulated flu B trees with 2,000–3,000 tips across four transmission scenarios. Colors indicate subtree size. Solid lines show the estimated mean $R_t$, with shaded bands representing the IQR. Dashed black lines denote the true underlying reproductive number.

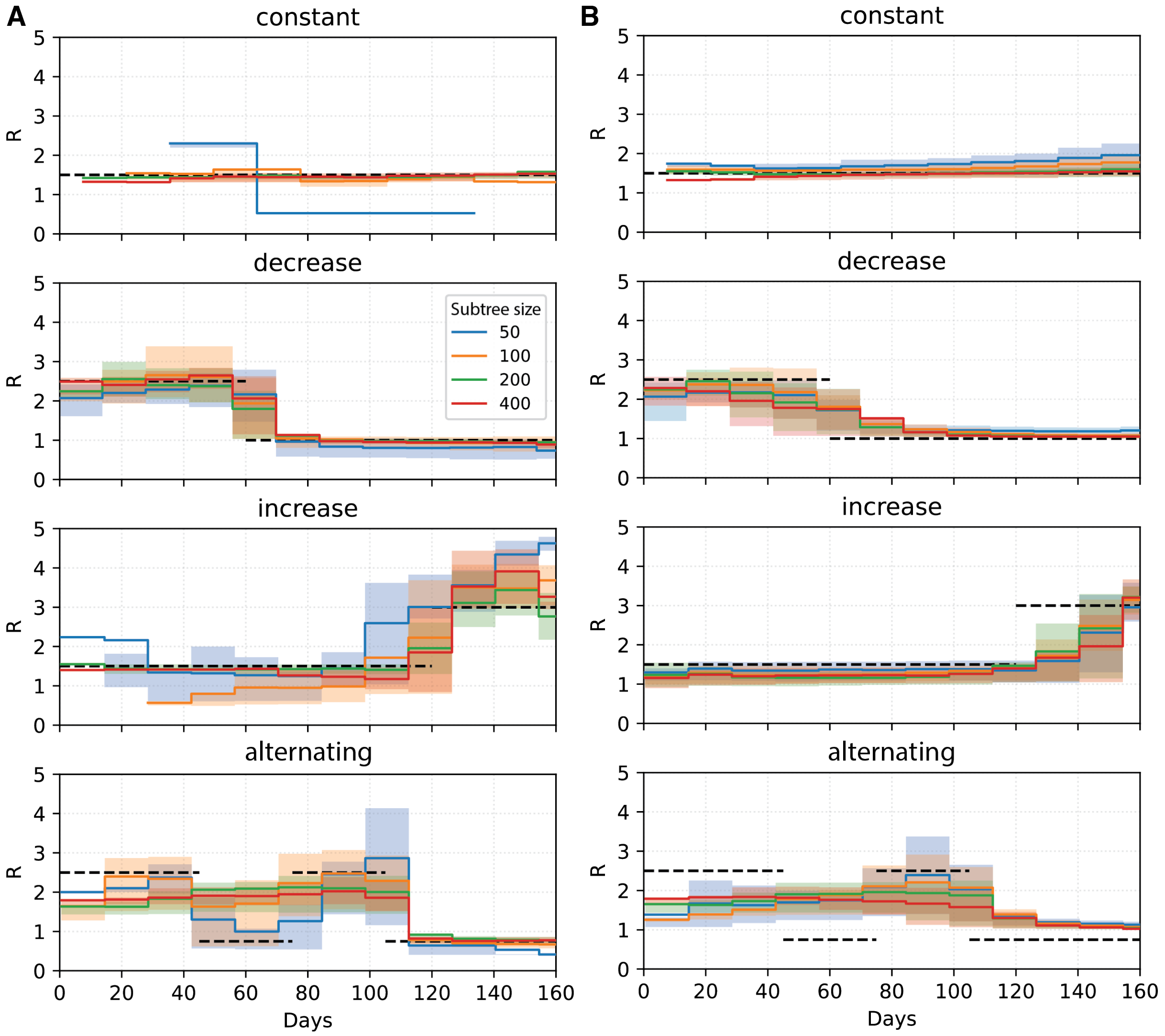


**Fig. S10.** Comparison of hierarchical (A) and baseline direct-averaging (B) subtree-based inference for estimating $R_t$ in three independently simulated SARS-CoV-2 trees with 2,000–3,000 tips across four transmission scenarios. Colors indicate subtree size. Solid lines show the estimated mean $R_t$, with shaded bands representing the IQR. Dashed black lines denote the true underlying reproductive number.

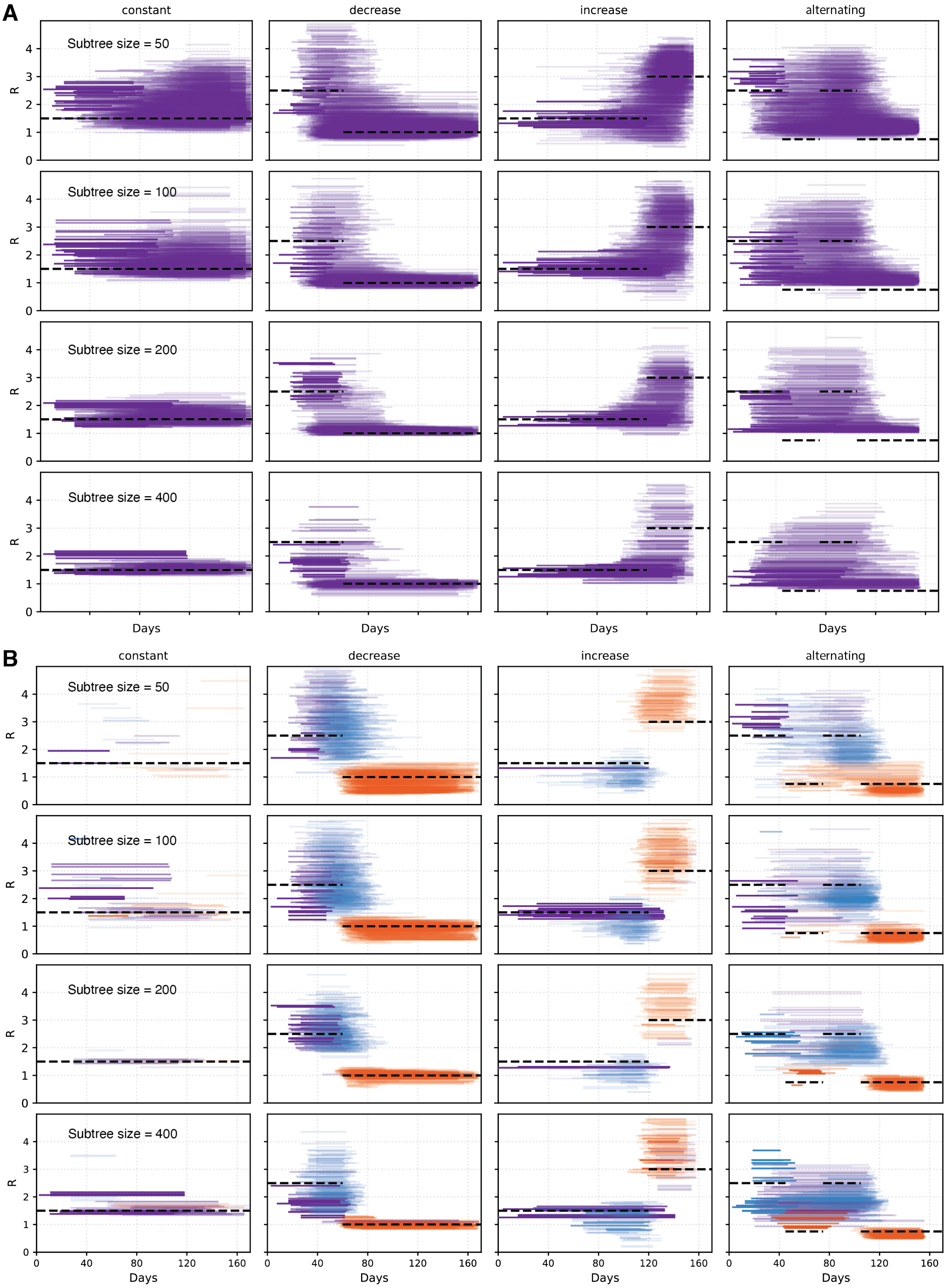


**Fig. S11.** Subtree-level reproductive number estimates for H3N2 simulated trees with 5,000–10,000 tips, using a baseline direct-averaging approach (A) and hierarchical inference (B) across transmission scenarios and subtree sizes.

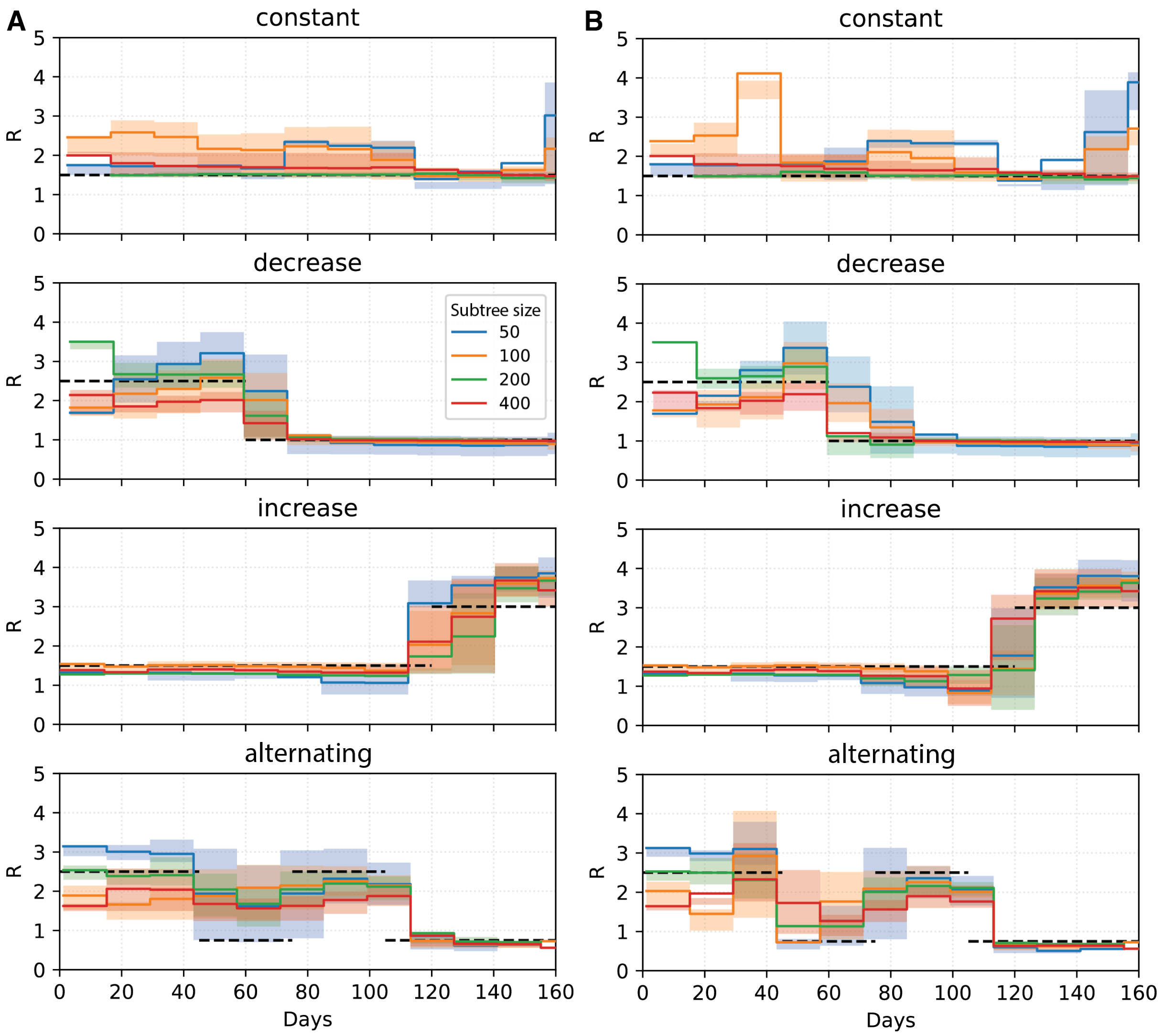


**Fig. S12.** Comparison of hierarchical subtree-based inference without (A) and with recency-based weighting (B) for estimating $R_t$ in three independently simulated H3N2 trees with 5,000–10,000 tips across four transmission scenarios. Colors indicate subtree size. Solid lines show the estimated mean $R_t$, with shaded bands representing the IQR. Dashed black lines denote the true underlying reproductive number.

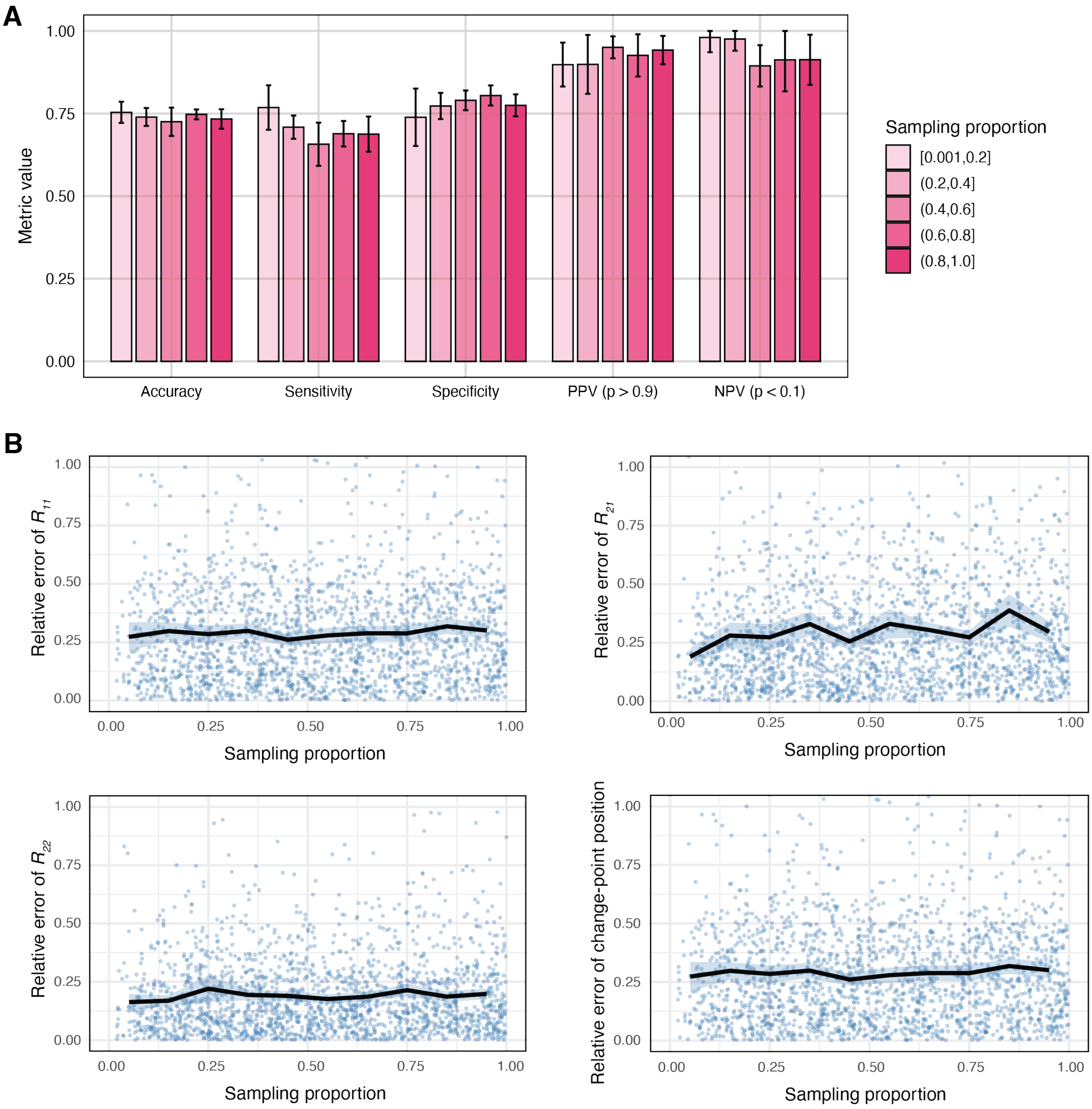


**Fig. S13.** Sensitivity of neural network model predictions to sampling proportion for models trained on 200-tip subtrees under the SARS-CoV-2 parameterization. (A) Classification performance across sampling-proportion intervals. Bars show mean values across testing subsets, with error bars indicating variability across 5 equal-sized splits of the testing dataset. For ACC, SN, and SP, predicted change-point probabilities were converted to binary labels using a threshold of 0.5. (B) Relative errors of regression outputs as a function of sampling proportion. Points represent individual predictions from the testing dataset. Solid black lines show the mean relative error computed within sampling-proportion intervals of width 0.1 spanning 0.001 to 1.0, plotted at the midpoint of each interval; shaded bands indicate 95% confidence intervals of the mean estimates.

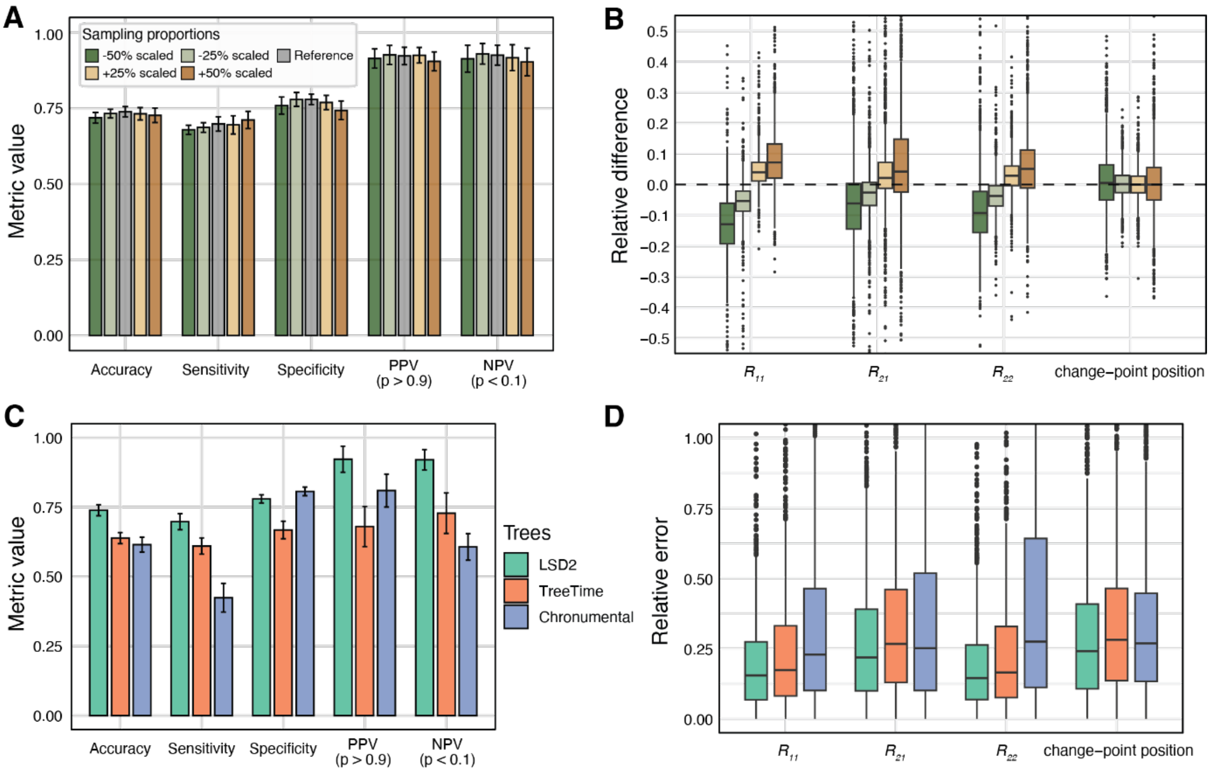


**Fig. S14.** Sensitivity analysis of neural network model predictions to sampling proportion and tree-dating method for models trained on 200-tip subtrees simulated under the SARS-CoV-2 parameterization. (A) Classification performance metrics under perturbed sampling proportions, shown relative to predictions obtained using the true sampling proportion. (B) Relative differences in regression outputs under perturbed sampling proportions. The horizontal dashed black line indicates zero relative difference, corresponding to predictions obtained using the true sampling proportion. (C) Classification performance metrics for trees dated using different toolkits. (D) Relative error distributions for regression outputs for trees dated using different methods. Bars show mean values across testing subsets, with error bars indicating variability across 10 equal-sized splits of the testing dataset. For accuracy (ACC), sensitivity (SN), and specificity (SP), predicted change-point probabilities were classified using a threshold of 0.5.

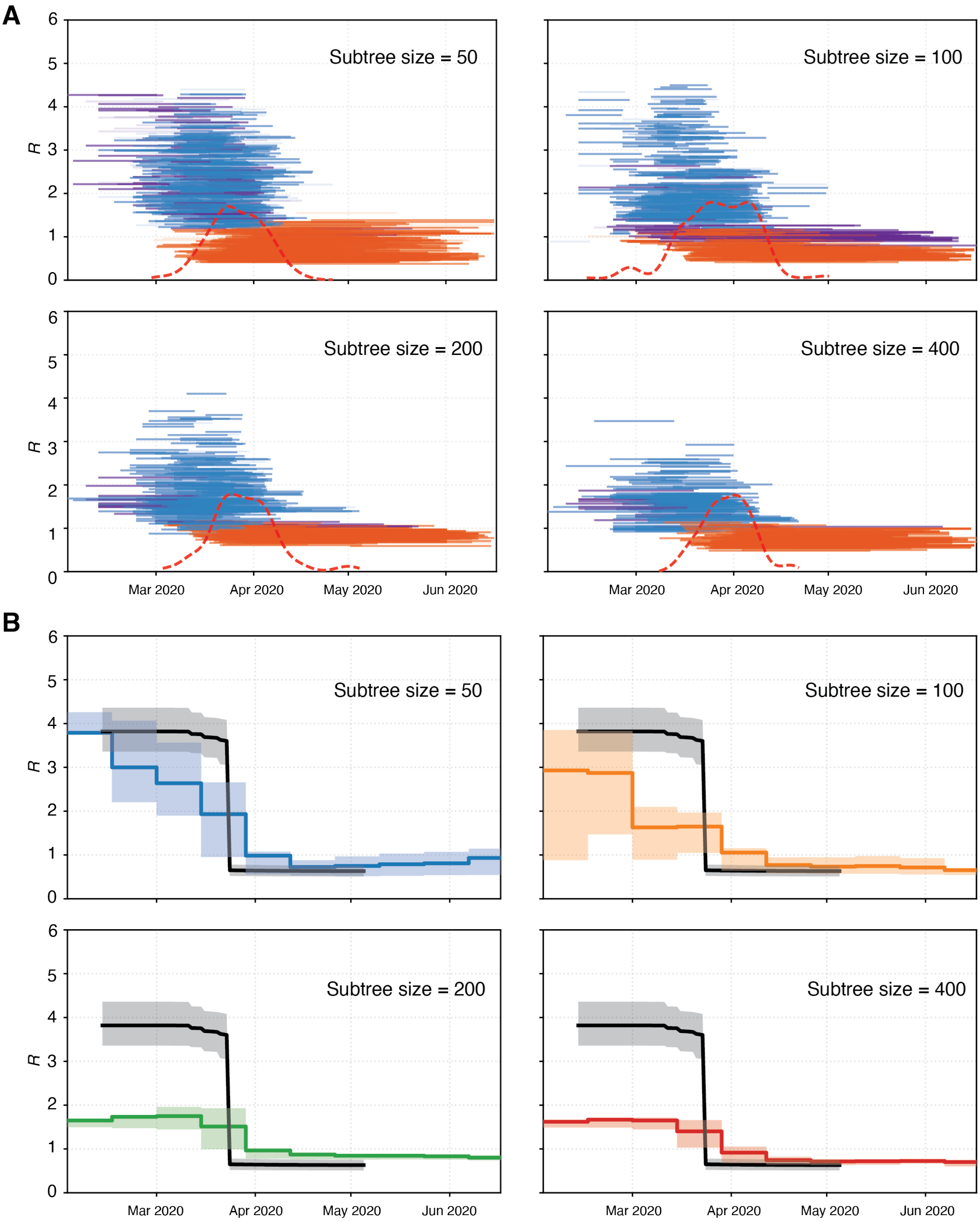


**Fig. S15.** Subtree-level $R$ (A) and $R_t$ (B) inferred by *PhyloRt* for the SARS-CoV-2 outbreak in the United Kingdom during early 2020 across different subtree sizes. Each horizontal segment in (A) represents an estimate from an individual subtree, coloured by $R_{NC}$, $R_{C1}$, and $R_{C2}$. The dashed red curve at the bottom shows a kernel-smoothed density of inferred change-point times across subtrees, normalized and scaled for visualization. In (B), *PhyloRt* estimates are shown with the IQR and compared with incidence-based surveillance estimates (black line, 95% confidence interval) (5).

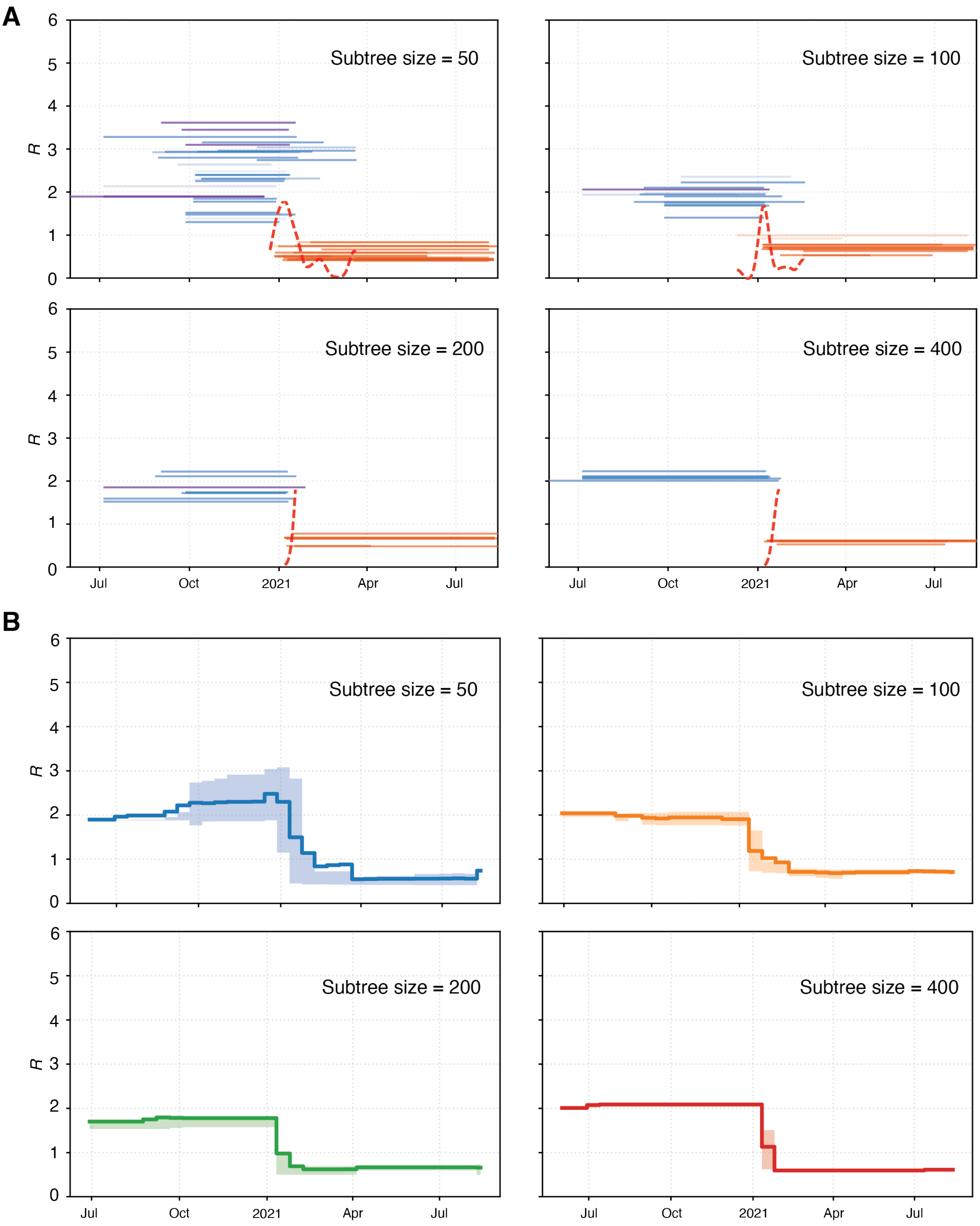


**Fig. S16.** Subtree-level $R$ (A) and $R_t$ (B) inferred by *PhyloRt* for influenza B/Victoria clade 3a1 circulating in China from 2020 to 2021 across different subtree sizes. Each horizontal segment in (A) represents an estimate from an individual subtree, coloured by $R_{NC}$, $R_{C1}$, and $R_{C2}$. The dashed red curve at the bottom shows a kernel-smoothed density of inferred change-point times across subtrees, normalized and scaled for visualization. In (B), *PhyloRt* estimates are shown with the IQR.

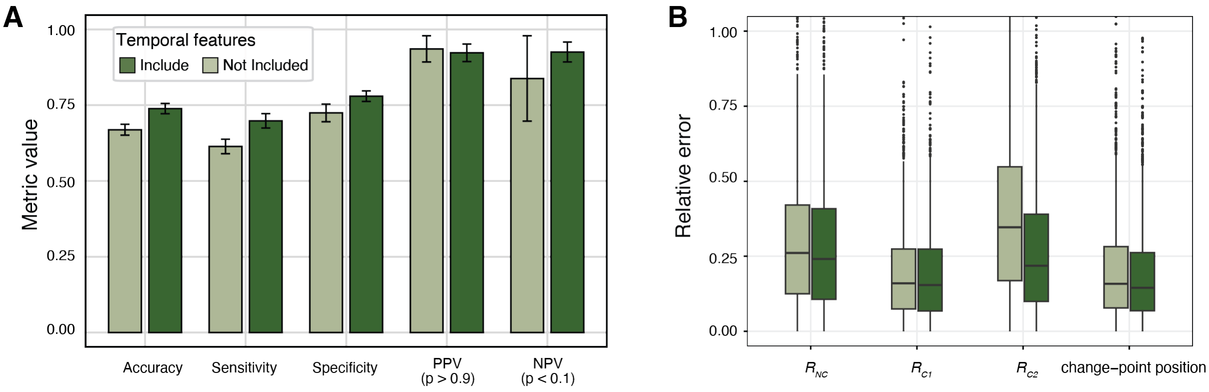


**Fig. S17.** Performance of neural network models trained on 200-tip subtrees simulated under the SARS-CoV-2 parameterization, with and without two tree-level temporal features that encode the subtree's relative position within the full tree. (A) Classification performance metrics with and without temporal features. Bars show mean values across testing subsets, with error bars indicating variability across 10 equal-sized splits of the testing dataset. For accuracy (ACC), sensitivity (SN), and specificity (SP), change-point probabilities were thresholded at 0.5. (B) Relative error distributions for regression outputs with and without temporal features.

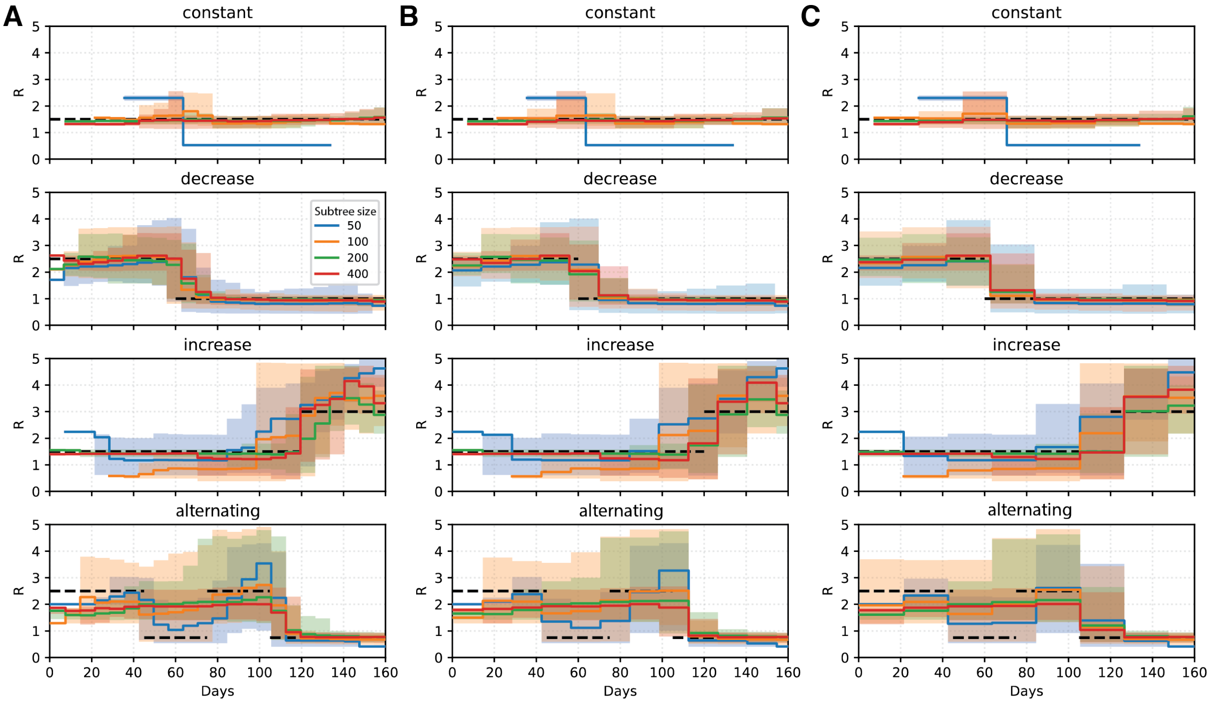


**Fig. S18.** Comparison of subtree-based inference using different time-bin sizes: 7 days (A), 14 days (B), and 21 days (C), for estimating $R_t$ in three independently simulated SARS-CoV-2 trees (2,000–3,000 tips) across four transmission scenarios. Colors indicate subtree size. Solid lines show the estimated mean $R_t$, with shaded bands representing the IQR. Dashed black lines denote the true underlying reproductive number.

**Table S1.** Parameter ranges used for simulations.

| Parameters | Name | Range | Range in large trees |
|---|---|---|---|
| $R$ | Reproduction number | (1, 6) | (**0.3**, 6)* |
| $1/\gamma$ | Infectious period | (1, 14) | (1, 14) |
| $\varepsilon$ | Incubation factor | (0.2, 5) | (0.2, 5) |
| $1/\varepsilon$ | Incubation period | (0.2, 70) | (0.2, 70) |
| $X_{ss}$ | Superspreading infectious ratio | (2, 40) | (2, 40) |
| $f_{ss}$ | Fraction of superspreaders | (0.01, 0.25) | (0.01, 0.25) |
| $t$ | Tree size | (200, 500) | (**1000**, **10000**) |
| $s$ | Sampling proportion | (0.01, 0.6) | (**0.001**, 1) |

**Values highlighted in bold indicate the adjusted boundary values.*

**Table S2.** Pathogen-specific birth–death models, genome lengths, and molecular clock rates used in simulations.

| Pathogen | Birth-death model | Clock rate* | Genome length | Ref. genome | Ref. for clock rate |
|---|---|---|---|---|---|
| SARS-CoV-2 | BDEISS | $8\times10^{-4} \pm 4\times10^{-4}$ | 29903 | Wuhan-Hu-1 | (6) |
| H3N2 | BDEI | $5\times10^{-3} \pm 1.02\times10^{-4}$ | 1737 | A/Wisconsin/67/2005 | (7) |
| H1N1 | BDEI | $4.4\times10^{-3} \pm 1.02\times10^{-4}$ | 1752 | A/California/07/2009 | (7) |
| Flu B | BDEI | $2\times10^{-3} \pm 2.12\times10^{-4}$ | 1885 | B/Brisbane/60/2008 | (8) |
| RSVA | BDEI | $6.47\times10^{-4} \pm 4.64\times10^{-5}$ | 15225 | hRSV/A/England/397/2017 | (9) |
| RSVB | BDEI | $7.76\times10^{-4} \pm 4.31\times10^{-5}$ | 15222 | hRSV/B/Australia/VIC-RCH056/2019 | (9) |

*mean ± standard deviation. Standard deviations were derived from published 95% confidence intervals under a normal approximation. For influenza viruses, simulations were based on the hemagglutinin (HA) segment only.

**Table S3.** Performance comparison of classification models across different subtree sizes (mean ± SD).

| Models | Subtree size | ACC (0.5)* | SN (0.5)* | SP (0.5)* | PPV (0.9)* | NPV (0.1)* |
|---|---|---|---|---|---|---|
| BDEI-H3N2 | 50 | 0.728±0.022 | 0.719±0.020 | 0.737±0.043 | 0.943±0.029 | 0.959±0.035 |
| | 100 | 0.749±0.030 | 0.762±0.029 | 0.736±0.037 | 0.966±0.020 | 0.958±0.030 |
| | 200 | 0.767±0.016 | 0.761±0.022 | 0.772±0.029 | 0.962±0.025 | 0.965±0.034 |
| | 400 | 0.776±0.019 | 0.761±0.027 | 0.792±0.026 | 0.927±0.042 | 0.957±0.022 |
| BDEI-flub | 50 | 0.728±0.016 | 0.697±0.021 | 0.759±0.023 | 0.922±0.062 | 0.961±0.039 |
| | 100 | 0.749±0.029 | 0.747±0.040 | 0.751±0.028 | 0.943±0.036 | 0.964±0.033 |
| | 200 | 0.776±0.019 | 0.778±0.019 | 0.773±0.028 | 0.957±0.025 | 0.961±0.032 |
| | 400 | 0.775±0.021 | 0.772±0.034 | 0.778±0.036 | 0.961±0.021 | 0.942±0.029 |
| BDEISS-COVID | 50 | 0.703±0.023 | 0.674±0.039 | 0.732±0.024 | 0.937±0.034 | 0.927±0.046 |
| | 100 | 0.710±0.022 | 0.700±0.021 | 0.718±0.035 | 0.932±0.038 | 0.926±0.054 |
| | 200 | 0.739±0.020 | 0.698±0.029 | 0.780±0.015 | 0.922±0.047 | 0.920±0.037 |
| | 400 | 0.758±0.026 | 0.729±0.031 | 0.786±0.038 | 0.953±0.036 | 0.945±0.025 |

** Values in parentheses indicate the decision thresholds used to compute each metric.*

**Table S4.** Performance comparison (mean relative error) of regression models.

| Models | Subtree *size* | $R_{NC}$ | $R_{C1}$ | $R_{C2}$ | *Change-point* |
|---|---|---|---|---|---|
| BDEI-H3N2 | 50 | 0.294 | 0.267 | 0.262 | 0.318 |
| | 100 | 0.224 | 0.233 | 0.212 | 0.286 |
| | 200 | 0.219 | 0.202 | 0.156 | 0.268 |
| | 400 | 0.168 | 0.179 | 0.128 | 0.235 |
| BDEI-flub | 50 | 0.307 | 0.289 | 0.255 | 0.298 |
| | 100 | 0.251 | 0.257 | 0.207 | 0.272 |
| | 200 | 0.195 | 0.222 | 0.170 | 0.252 |
| | 400 | 0.174 | 0.216 | 0.141 | 0.228 |
| BDEISS-COVID | 50 | 0.317 | 0.367 | 0.291 | 0.328 |
| | 100 | 0.279 | 0.326 | 0.250 | 0.305 |
| | 200 | 0.215 | 0.298 | 0.192 | 0.290 |
| | 400 | 0.194 | 0.265 | 0.149 | 0.262 |

**Table S5.** Time-normalized mean absolute error (MAE) of *R* for hierarchical and baseline direct-averaging inference across structured time-varying transmission scenarios in H3N2 based on trees with tip size from 2000-3000.

| Pathogen | *Subtree size* | *Direct averaging* | | | | *Hierarchical* | | | |
|---|---|---|---|---|---|---|---|---|---|
| | | constant | *decrease* | *increase* | *zigzag* | constant | *decrease* | *increase* | *zigzag* |
| H3N2 | 50 | 0.462 | **0.239** | **0.407** | **0.681** | 0.655 | 0.283 | 0.320 | 0.498 |
| | 100 | 0.260 | 0.278 | **0.407** | 0.752 | 0.328 | 0.250 | 0.367 | **0.389** |
| | 200 | 0.191 | 0.324 | 0.487 | 0.745 | 0.062 | 0.196 | **0.145** | 0.519 |
| | 400 | **0.109** | 0.382 | 0.470 | 0.763 | **0.040** | **0.176** | 0.366 | 0.610 |
| Flu B | 50 | 0.540 | **0.244** | 0.600 | 0.738 | 0.513 | 0.321 | **0.434** | **0.512** |
| | 100 | 0.203 | 0.278 | 0.572 | 0.756 | 0.187 | 0.166 | 0.455 | 0.519 |
| | 200 | 0.264 | 0.389 | **0.553** | 0.835 | 0.230 | 0.155 | 0.472 | 0.589 |
| | 400 | **0.037** | 0.345 | 0.597 | **0.656** | **0.061** | **0.077** | 0.570 | 0.591 |
| SARS-CoV-2 | 50 | 0.275 | 0.315 | **0.286** | 0.675 | 0.925 | 0.287 | 0.701 | 0.499 |
| | 100 | 0.141 | 0.232 | 0.331 | 0.735 | 0.123 | 0.152 | 0.585 | **0.392** |
| | 200 | **0.044** | **0.227** | 0.366 | 0.679 | 0.074 | **0.139** | **0.152** | 0.570 |
| | 400 | 0.059 | 0.290 | 0.410 | **0.641** | **0.066** | 0.154 | 0.276 | 0.508 |

*Bold values indicate the lowest MAE for each transmission scenario within a given tree-size category.*

**Table S6.** Time-normalized mean absolute error (MAE) and coverage (in parentheses) for hierarchical and baseline direct-averaging inference across structured time-varying transmission scenarios in H3N2 based on trees with 5000-10000 tip size.

| Recency weight | *Subtree size* | *Direct averaging* | | | | *Hierarchical* | | | |
|---|---|---|---|---|---|---|---|---|---|
| | | constant | *decrease* | *increase* | *zigzag* | constant | *decrease* | *increase* | *zigzag* |
| 0 | 50 | 0.536 | 0.277 | **0.157** | **0.603** | 0.449 | 0.335 | 0.486 | 0.563 |
| | 100 | 0.425 | **0.260** | 0.164 | 0.687 | 0.580 | 0.237 | **0.195** | 0.572 |
| | 200 | 0.174 | 0.272 | 0.260 | 0.762 | **0.024** | **0.189** | 0.328 | **0.441** |
| | 400 | **0.123** | 0.312 | 0.271 | 0.789 | 0.184 | 0.214 | 0.256 | 0.561 |
| 4 | 50 | - | - | - | - | 0.678 | 0.412 | 0.414 | 0.436 |
| | 100 | - | - | - | - | 0.694 | 0.326 | **0.218** | 0.438 |
| | 200 | - | - | - | - | **0.040** | **0.163** | 0.276 | **0.307** |
| | 400 | - | - | - | - | 0.180 | 0.176 | 0.350 | 0.511 |

*Bold values indicate the lowest MAE for each transmission scenario within a given tree-size category.*
